# An open-label randomized, controlled trial of the effect of lopinavir/ritonavir, lopinavir/ritonavir plus IFN-β-1a and hydroxychloroquine in hospitalized patients with COVID-19 – Final results from the DisCoVeRy trial

**DOI:** 10.1101/2022.02.16.22271064

**Authors:** Florence Ader, Nathan Peiffer-Smadja, Julien Poissy, Maude Bouscambert-Duchamp, Drifa Belhadi, Alpha Diallo, Christelle Delmas, Juliette Saillard, Aline Dechanet, Noémie Mercier, Axelle Dupont, Toni Alfaiate, François-Xavier Lescure, François Raffi, François Goehringer, Antoine Kimmoun, Stéphane Jaureguiberry, Jean Reignier, Saad Nseir, François Danion, Raphael Clere-Jehl, Kévin Bouiller, Jean-Christophe Navellou, Violaine Tolsma, André Cabie, Clément Dubost, Johan Courjon, Sylvie Leroy, Joy Mootien, Rostane Gaci, Bruno Mourvillier, Emmanuel Faure, Valérie Pourcher, Sébastien Gallien, Odile Launay, Karine Lacombe, Jean-Philippe Lanoix, Alain Makinson, Guillaume Martin-Blondel, Lila Bouadma, Elisabeth Botelho-Nevers, Amandine Gagneux-Brunon, Olivier Epaulard, Lionel Piroth, Florent Wallet, Jean-Christophe Richard, Jean Reuter, Thérèse Staub, Bruno Lina, Marion Noret, Claire Andrejak, Minh Patrick Lê, Gilles Peytavin, Maya Hites, Dominique Costagliola, Yazdan Yazdanpanah, Charles Burdet, France Mentre, the DisCoVeRy study group

## Abstract

**Objectives:** We evaluated the clinical, virological and safety outcomes of lopinavir/ritonavir, lopinavir/ritonavir-interferon (IFN)-β-1a, hydroxychloroquine or remdesivir in comparison to standard of care (control) in COVID-19 inpatients requiring oxygen and/or ventilatory support. While preliminary results were previously published, we present here the final results, following completion of the data monitoring.

**Methods:** We conducted a phase 3 multi-centre open-label, randomized 1:1:1:1:1, adaptive, controlled trial (DisCoVeRy), add-on trial to Solidarity (NCT04315948, EudraCT2020-000936-23). The primary outcome was the clinical status at day 15, measured by the WHO 7-point ordinal scale. Secondary outcomes included SARS-CoV-2 quantification in respiratory specimens, pharmacokinetic and safety analyses. We report the results for the lopinavir/ritonavir-containing arms and for the hydroxychloroquine arm, which were stopped prematurely.

**Results:** The intention-to-treat population included 593 participants (lopinavir/ritonavir, n=147; lopinavir/ritonavir-IFN-β-1a, n=147; hydroxychloroquine, n=150; control, n=149), among whom 421 (71.0%) were male, the median age was 64 years (IQR, 54-71) and 214 (36.1%) had a severe disease. The day 15 clinical status was not improved with investigational treatments: lopinavir/ritonavir *versus* control, adjusted odds ratio (aOR) 0.82, (95% confidence interval [CI] 0.54-1.25, P=0.36); lopinavir/ritonavir-IFN-β-1a *versus* control, aOR 0.69 (95%CI 0.45-1.05, P=0.08); hydroxychloroquine *versus* control, aOR 0.94 (95%CI 0.62-1.41, P=0.76). No significant effect of investigational treatment was observed on SARS-CoV-2 clearance. Trough plasma concentrations of lopinavir and ritonavir were higher than those expected, while those of hydroxychloroquine were those expected with the dosing regimen. The occurrence of Serious Adverse Events was significantly higher in participants allocated to the lopinavir/ritonavir-containing arms.

**Conclusion:** In adults hospitalized for COVID-19, lopinavir/ritonavir, lopinavir/ritonavir-IFN-ß-1a and hydroxychloroquine did not improve the clinical status at day 15, nor SARS-CoV-2 clearance in respiratory tract specimens.

## Introduction

Worldwide research efforts against SARS-CoV-2 initially focused on repurposed drugs that showed broad-spectrum antiviral activity against coronaviruses (1,2). Lopinavir/ritonavir (3,4), type I interferon (IFN) (5–7), hydroxychloroquine (8–10), and remdesivir (11) were among the first investigational treatments to be tested on the basis of their *in vitro* activity against SARS-CoV-2.

The DisCoVeRy trial is a European randomized controlled trial evaluating the clinical and the virological efficacy, as well as the safety, of lopinavir/ritonavir, lopinavir/ritonavir plus IFN-β-1a, hydroxychloroquine, and remdesivir as compared with standard of care in adults hospitalized for COVID-19 (12). As an add-on trial to the international Solidarity trial sponsored by the World Health Organization (WHO), it has contributed to data acquisition on in-hospital mortality, need for mechanical ventilation and time to hospital discharge. Interim analyses of these variables concluded to futility, leading to discontinuation of three treatment arms while inclusions continued in the remdesivir arm (13). The DisCoVeRy trial was designed to further document clinical outcomes, virological kinetics, treatment pharmacokinetics and related safety data. We report here the final results for the lopinavir/ritonavir, lopinavir/ritonavir plus IFN-β-1a and hydroxychloroquine arms, after completion of the data monitoring. Preliminary results were previously published (14). Results for the remdesivir arm have been presented separately (15).

## Methods

### Trial design and oversight

DisCoVeRy is a phase 3 open-label, adaptive, multicenter, randomized, superiority-controlled trial which evaluates the efficacy and safety of repurposed drugs in adults hospitalized for COVID-19. Sponsored by the Institut national de la santé et de la recherche médicale (Inserm, France), the trial was approved by the Ethics Committee (CPP Ile-de-France-III, approval #20.03.06.51744). Written informed consent was obtained from all included participants or their legal representative, when unable to consent. The trial was conducted in accordance with the Declaration of Helsinki and national laws and regulations and declared on the clinicatrials.gov registry (NCT 04315948) and on the European Clinical Trials Database (2020-000936-23).

### Study population

Eligible participants were adults (≥ 18-year-old) hospitalized with a PCR-positive (< 72 hours) SARS-CoV-2 infection and pulmonary rales or crackles with a peripheral oxygen saturation ≤ 94% or requiring supplemental oxygen. Inclusion and exclusion criteria are presented in the Supplementary Appendix.

### Interventions and randomization

Participants were randomly assigned to treatment arms in a 1:1:1:1 ratio, through computer-generated blocks of various sizes and stratification by administrative region and severity of disease at enrolment (moderate: hospitalized participants not requiring oxygen or receiving low-flow supplemental oxygen; severe: hospitalized participants requiring non-invasive ventilation or high-flow oxygen devices, invasive mechanical ventilation or extracorporeal membrane oxygenation (ECMO)). Randomization was implemented in the electronic Case Report Form to ensure appropriate allocation concealment. Investigational arms were standard of care (SoC, control), SoC plus lopinavir/ritonavir (400 mg lopinavir and 100 mg ritonavir orally twice on day for 14 days (3,16)), SoC plus lopinavir/ritonavir plus IFN-ß-1a (44 μg of subcutaneous IFN-ß-1a on days 1, 3, and 6), SoC plus hydroxychloroquine (400 mg orally, twice on day 1 as a loading dose followed by 400 mg once daily for 9 days) (17). Supportive treatments corticosteroids, anticoagulants or immunomodulatory agents were allowed except antivirals. Enrolment in another investigative trial was not allowed.

### Clinical and laboratory monitoring

Participants were assessed at days 3, 5, 8, 11, 15±2 and 29±3 while hospitalized. If discharge occurred before day 15, face-to-face visits were set up for days 15±2 and 29±3, for efficacy and safety evaluations. Clinical data, concomitant medications, adverse events (AEs) and measurements for safety biological data (blood cell counts, serum creatinine and liver aminotransferases) were collected. Nasopharyngeal swab (NPS) and lower respiratory tract (LRT) specimens were collected for SARS-CoV-2 RNA quantification. For lopinavir and ritonavir, trough plasma concentrations were obtained at days 1 and 3, 12h (±2h) after the last administration and for hydroxychloroquine, at day 1 12h (±2h) and at day 3 24h (±4h) after the last administration.

### Outcomes measures

The primary outcome measure was the clinical status at day 15 as measured on the 7-point ordinal scale of the WHO Master Protocol (v3.0, March 3, 2020): 1. Not hospitalized, no limitation on activities; 2. Not hospitalized, limitation on activities; 3. Hospitalized, not requiring supplemental oxygen; 4. Hospitalized, requiring supplemental oxygen; 5. Hospitalized, on non-invasive ventilation or high flow oxygen devices; 6. Hospitalized, on invasive mechanical ventilation or ECMO; 7. Death.

Secondary efficacy outcome measures were the clinical status at day 29 and the time to an improvement of 2 categories as measured on the 7-point ordinal scale or hospital discharge until day 29, the time to National Early Warning Score 2 (NEWS2) ≤2 or hospital discharge until day 29, the time to hospital discharge until day 29, oxygenation- and ventilator-free days until day 29, in-hospital, 29-day and 3-month mortality, and the SARS-CoV-2 detection and quantitative normalized viral loads. Trough plasma concentrations of lopinavir, ritonavir and hydroxychloroquine were measured at days 1 and 3.

Secondary safety outcomes included the cumulative incidence of any grade 3 or 4 AE, or of any serious adverse event (SAE, according to the DAIDS Table for Grading the Severity of Adult and Paediatric Adverse Events, v2.1, July 2017) and the proportion of patients with a premature suspension or discontinuation for any reason of the investigational treatments.

### Virological methods

Determination of normalized viral load blinded to treatment arm was performed on NPS and LRT specimens by RNA extraction on the EMAG® platform (bioMerieux, Marcy-l’Étoile, France). The SARS-CoV-2 load was measured by quantitative RT-PCR, according to a scale of calibrated in-house plasmid, using the RT-PCR RdRp-IP4 developed by the Institut Pasteur (Paris, France) (18). The amplification protocol was developed using QuantStudio 5 rtPCR Systems (Thermo Fisher Scientific, Waltham, Massachusetts, USA). The number of cells in sample (quality criteria for NPS and normalization tool for viral load determination) was checked using the CELL Control r-gene® kit (Argene-BioMérieux, Marcy-l’Étoile, France). If cell quantification was below 500 cells/reaction, the quality of the sample was considered too low to be measured. We computed a normalized SARS-CoV-2 load by dividing the viral load by the number of cells. All viral loads strictly below 1 log_10_ RNA copies/10 000 cells were considered under the limit of detection and were reported as negative.

### Pharmacological methods

Plasma concentrations of lopinavir, ritonavir and hydroxychloroquine were determined using liquid chromatography coupled with tandem mass spectrometry (19,20). The limits of quantification were 30 ng/mL for lopinavir and ritonavir, and 10 ng/mL for hydroxychloroquine.

### Sample size calculation

The sample size was determined assuming the following scenario under SoC for each item of the ordinal scale at day 15: 1, 42%; 2, 38%; 3, 8%; 4, 7%; 5, 2%; 6, 1%; 7, 2%. At the time of the trial design, there was a significant uncertainty with these assumptions. We powered the study for an odds ratio of 1.5 (an odds ratio higher than 1 indicates superiority of the experimental treatment over the control for each ordinal scale category), with 90% power and using an overall two-sided type I error rate of 0.05. Adjusting for multiplicity of 4 pairwise comparisons with the control arm in a 5-arm setting, the two-sided false positive error rate would be 0.0125. We determined that the inclusion of 620 patients in each treatment arm was required.

### Statistical and interim analyses

An independent data safety and monitoring board (DSMB) externally reviewed the trial data periodically. Based on interim analyses (see Supplementary Appendix), enrolment in the hydroxychloroquine arm was prematurely stopped on June 17^th^, and enrolment in lopinavir-containing arms was stopped on June 29^th^ 2020.

For the 7-point ordinal scale, data were analyzed using a proportional odds model, which assumes a common odds ratio between the 7 points of the ordinal scale. All analyses were stratified by severity at randomization, and adjusted effect measures are reported. Full statistical methods are presented in Supplementary Appendix.

## Results

### Patient’s characteristics at baseline

Between March 22^nd^ and June 29^th^ 2020, 603 participants were randomized across 30 sites in France and 2 in Luxembourg; 593 were evaluable for analysis (Supplementary Figure S1): control arm, n=149; lopinavir/ritonavir arm, n=147; lopinavir/ritonavir plus IFN-ß-1a arm, n=147; hydroxychloroquine arm, n=150. Participants’ baseline characteristics are presented in Table 1. Participants were mostly male (n=421, 71.0 %), median age was 64 years (IQR, 54-71). The median time from symptoms onset to randomization was 9 days (IQR, 7-12). The most frequent underlying conditions were obesity (n=169, 28.7%), chronic cardiac disease (n=155, 26.2 %) and diabetes mellitus (n=134, 22. 7%). At baseline, severe disease accounted for 214 (36.1%) participants. Concomitant treatments are listed in Supplementary Table 1.

**Table 1.** Baseline characteristics of patients included in the intention to treat population of the present analysis of DisCoVeRy trial. NPS, Nasopharyngeal swabs; LRT, Lower respiratory tract. * denotes variables with missing data. Data on chronic cardiac disease, chronic pulmonary disease, mild liver disease, chronic neurological disorder, active cancer and diabetes mellitus were missing for 2 patients; data on chronic kidney disease were missing for 3 patients; data on auto-inflammatory disease were missing for 1 patient; data on obesity were missing for 5 patients; data on smoking status were missing for 29 patients; data on the time from symptoms onset to randomization were missing for 7 patients; data on BMI were missing for 80 patients; data on randomization site were missing for 1 patient; data on viral load from NPS were missing for 236 patients; data on viral load from LRT specimens were missing for 538 patients; data for lymphocyte count were missing for 93 patients; data for neutrophil count were missing for 140 patients; data on creatinine were missing for 9 patients; data on AST/SGOT were missing for 40 patients; data on ALT/SGPT were missing for 37 patients; data on CRP were missing for 133 patients; data on D-Dimers were missing for 304 patients; data on PCT were missing for 362 patients; data on ferritin were missing for 426 patients. ** moderate disease: hospitalized participants receiving low-flow supplemental oxygen or not requiring oxygen; severe disease: hospitalized participants requiring non-invasive ventilation or high-flow oxygen devices, invasive mechanical ventilation or ECMO

|  | **Overall**<br>**(N=593)** | **Control (N=149)** | **Lopinavir/ritonavir (L/r)**<br>**(N=147)** | **Lopinavir/Ritonavir +**<br>**Interferon β-1a (L/r + IFN)**<br>**(N=147)** | **Hydroxychloroquine (HCQ)**<br>**(N=150)** |
| --- | --- | --- | --- | --- | --- |
| **Median age — yr [IQR]** | 64 [54-71] | 62 [52-71] | 62 [55-71] | 64 [53-71] | 66 [55-71] |
| **Median BMI — kg/m<sup>2</sup> [IQR]** | 28 [25-32] | 28 [25-33] | 28 [26-31] | 28 [26-32] | 28 [25-31] |
| **Male sex — no. (%)** | 421 (71.0%) | 106 (71.1%) | 107 (72.8%) | 103 (70.1%) | 105 (70.0%) |
| **Coexisting condition\* — no. (%)** |  |  |  |  |  |
| - Chronic cardiac disease | 155 (26.2%) | 40 (26.8%) | 36 (24.5%) | 36 (24.8%) | 43 (28.7%) |
| - Chronic pulmonary disease | 90 (15.2%) | 32 (21.5%) | 20 (13.6%) | 19 (13.1%) | 19 (12.7%) |
| - Chronic kidney disease (stage 1 to 3) | 26 (4.4%) | 8 (5.4%) | 2 (1.4%) | 6 (4.1%) | 10 (6.7%) |
| - Mild liver disease | 13 (2.2%) | 6 (4.0%) | 3 (2.0%) | 0 (0.0%) | 4 (2.7%) |
| - Chronic neurological disorder (including dementia) | 23 (3.9%) | 7 (4.7%) | 5 (3.4%) | 3 (2.1%) | 8 (5.3%) |
| - Active cancer | 36 (6.1%) | 10 (6.7%) | 8 (5.4%) | 6 (4.1%) | 12 (8.0%) |
| - Auto-inflammatory disease | 26 (4.4%) | 8 (5.4%) | 4 (2.7%) | 9 (6.2%) | 5 (3.3%) |

|  | **Overall**<br>**(N=593)** | **Control (N=149)** | **Lopinavir/ritonavir (L/r)**<br>**(N=147)** | **Lopinavir/Ritonavir +**<br>**Interferon β-1a (L/r + IFN)**<br>**(N=147)** | **Hydroxychloroquine (HCQ)**<br>**(N=150)** |
| --- | --- | --- | --- | --- | --- |
| - Obesity | 169 (28.7%) | 46 (31.3%) | 36 (24.5%) | 41 (28.1%) | 46 (31.1%) |
| - Diabetes mellitus | 134 (22.7%) | 36 (24.2%) | 36 (24.5%) | 28 (19.3%) | 34 (22.7%) |
| - Current smoker | 17 (3.0%) | 5 (3.5%) | 4 (2.9%) | 4 (2.8%) | 4 (2.9%) |
| **Median time from symptom onset to**<br>**randomization\* — days [IQR]** | 9.0 [7.0-12.0] | 10.0 [7.0-12.0] | 10.0 [7.0-13.0] | 10.0 [7.0-12.0] | 8.0 [7.0-11.0] |
| **Baseline severity of COVID-19\*\* — no. (%)** |  |  |  |  |  |
| - Moderate | 379 (63.9%) | 95 (63.8%) | 95 (64.6%) | 93 (63.3%) | 96 (64.0%) |
| - Severe | 214 (36.1%) | 54 (36.2%) | 52 (35.4%) | 54 (36.7%) | 54 (36.0%) |
| **Randomization site\* — no. (%)** |  |  |  |  |  |
| - ICU | 255 (43.1%) | 64 (43.0%) | 66 (44.9%) | 63 (43.2%) | 62 (41.3%) |
| - Conventional unit (e.g. : infectious disease<br>unit, internal medicine, pneumology) | 337 (56.9%) | 85 (57.0%) | 81 (55.1%) | 83 (56.8%) | 88 (58.7%) |
| **7-point ordinal scale at baseline — no. (%)** |  |  |  |  |  |
| - 3. Hospitalized, not requiring supplemental<br>oxygen | 21 (3.5%) | 5 (3.4%) | 4 (2.7%) | 7 (4.8%) | 5 (3.3%) |
| - 4. Hospitalized, requiring supplemental<br>oxygen | 354 (59.7%) | 87 (58.4%) | 89 (60.5%) | 87 (59.2%) | 91 (60.7%) |
| - 5. Hospitalized, on non-invasive ventilation<br>or high flow oxygen devices | 62 (10.5%) | 22 (14.8%) | 14 (9.5%) | 12 (8.2%) | 14 (9.3%) |
| - 6. Hospitalized, on invasive mechanical<br>ventilation or ECMO | 156 (26.3%) | 35 (23.5%) | 40 (27.2%) | 41 (27.9%) | 40 (26.7%) |
| **Median NEWS-2 at baseline\*, median**<br>**[IQR]** | 9.0 [7.0-12.0] | 9.0 [7.0-12.0] | 9.0 [7.0-11.0] | 10.0 [7.0-12.0] | 9.0 [6.0-11.0] |
| **Median viral load at baseline, median [IQR]** |  |  |  |  |  |
| - on NPS (log10 copies/10 000 cells) | 2.4 [1.0-3.7]<br>n=357) | 2.5 [1.2-4.0]<br>(n=87) | 2.5 [0.7-3.6]<br>(n=91) | 2.6 [1.0-3.8]<br>(n=81) | 2.1 [0.7-3.4]<br>(n=98) |
| - on LRT specimens (log10 copies/10 000<br>cells) | 4.3 [2.7-4.9]<br>n=55) | 3.8 [2.7-4.5]<br>(n=13) | 4.5 [3.2-4.8]<br>(n=14) | 3.5 [1.0-4.8]<br>(n=10) | 4.4 [3.2-5.5]<br>(n=18) |
| **Biological data at baseline\*, median [IQR]** |  |  |  |  |  |
| - Minimal lymphocytes count (G/L) | 0.9 [0.6-1.2] | 0.9 [0.6-1.3] | 0.8 [0.6-1.2] | 0.9 [0.7-1.3] | 0.9 [0.6-1.1] |
| - Maximal neutrophils count (G/L) | 5.8 [4.1-7.8] | 5.7 [4.1-7.8] | 6.3 [4.4-7.8] | 5.7 [4.0-8.2] | 5.5 [3.7-7.7] |
| - Maximal plasma creatinine (μmol/L) | 74.0 [61.5-91.0] | 73.5 [60.0-93.0] | 73.0 [62.0-88.0] | 77.0 [62.0-91.0] | 73.0 [62.0-91.0] |
| - Maximal SGOT (U/L) | 49.0 [35.0-73.0] | 52.0 [37.0-73.0] | 47.0 [34.0-65.0] | 48.5 [36.0-70.0] | 55.0 [34.0-81.0] |
| - Maximal SGPT (U/L) | 37.0 [25.0-62.5] | 40.5 [25.0-61.5] | 34.0 [23.0-59.0] | 37.0 [25.0-60.0] | 42.0 [26.0-67.5] |
| - Maximal plasma C-reactive protein (mg/L) | 118.0 [70.0-184.0] | 130.5 [73.0-191.5] | 123.0 [75.0-187.0] | 105.0 [66.0-167.0] | 115.0 [70.0-187.0] |
| - Maximal plasma D-dimers (μg/L) | 1060.0 [640.0-1860.0] | 1117.5 [688.0-2000.0] | 1054.5 [626.5-1938.5] | 970.0 [560.0-1850.0] | 1102.5 [545.0-1780.5] |
| - Maximal procalcitonin (ng/mL) | 0.2 [0.1-0.9] | 0.3 [0.1-1.1] | 0.2 [0.1-0.6] | 0.3 [0.1-0.9] | 0.3 [0.1-0.8] |
| - Maximal ferritin (mg/L) | 522.0 [2.0-1363.0] | 162.0 [2.0-1171.0] | 637.0 [2.0-1301.0] | 761.0 [3.0-1344.0] | 377.0 [2.0-1610.0] |

### Primary endpoint

The distribution of the 7-point ordinal scale at day 15 is presented in Figure 1 and Table 2. Adjusted OR for clinical improvement (aOR) were not in favor of investigational treatments (*i.e*., below 1): lopinavir/ritonavir *versus* control, aOR 0.82 (95% confidence interval [CI] 0.54-1.25, P=0.36); lopinavir/ritonavir plus IFN-ß-1a *vs*. control, aOR 0.69 (95%CI 0.45-1.05, P=0.08); hydroxychloroquine *vs*. control, aOR 0.94 (95%CI 0.62-1.41, P=0.76).

**Table 2.**
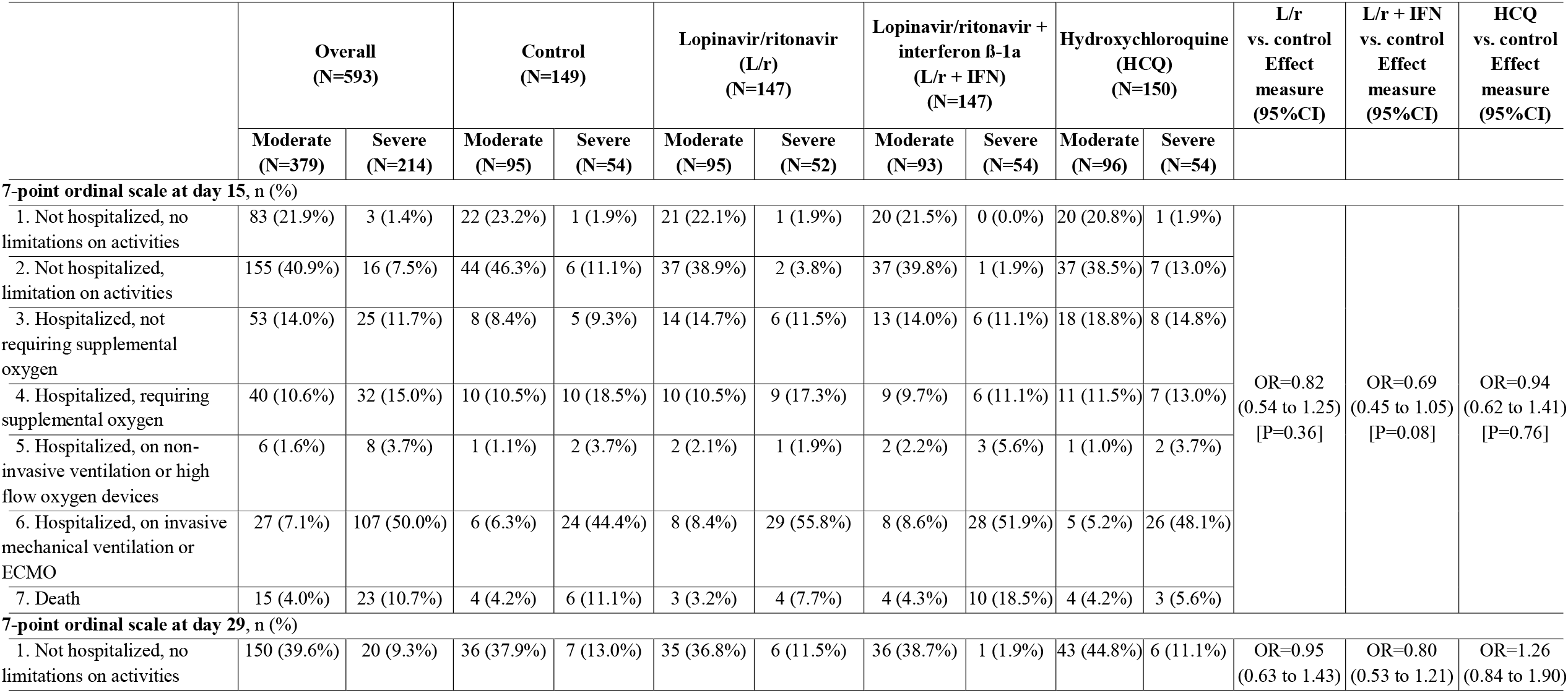
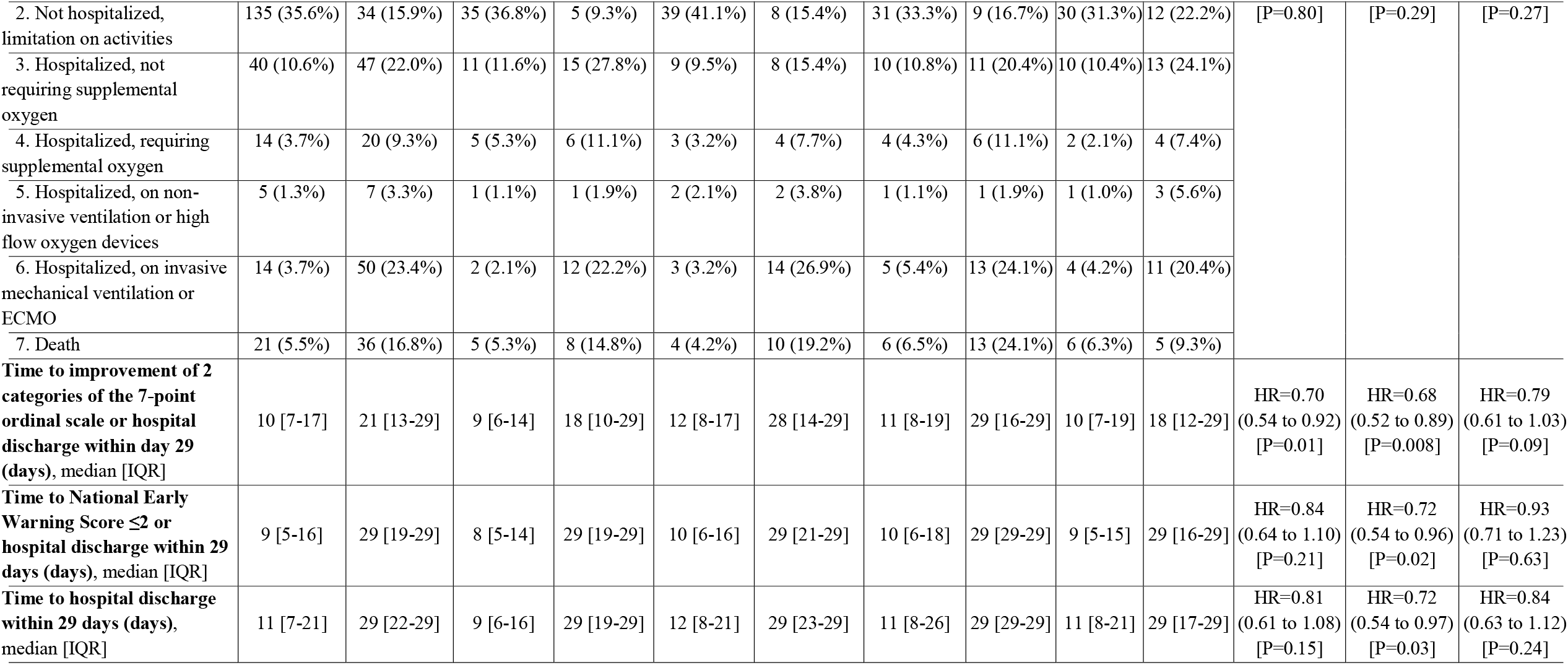
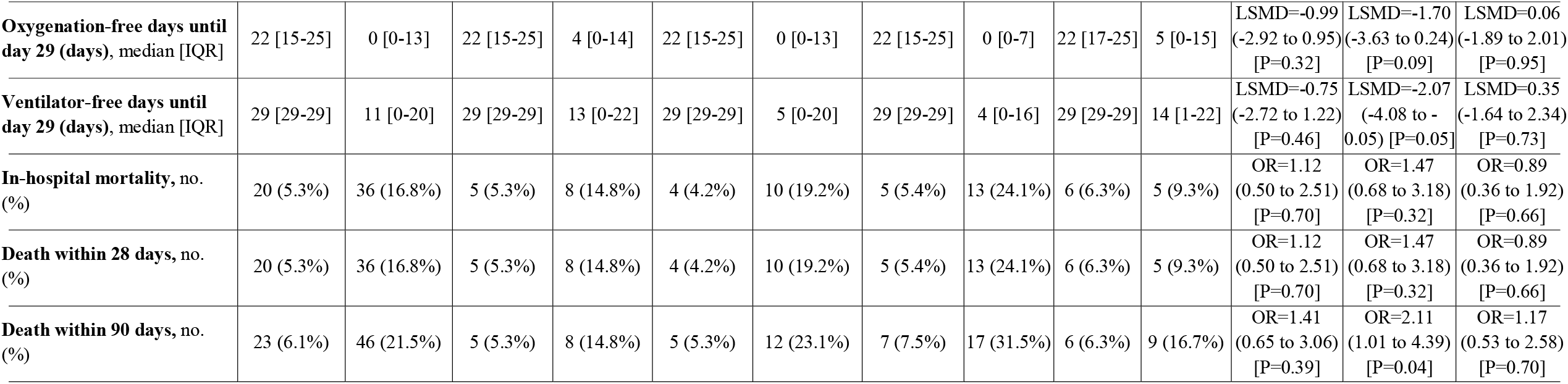
Primary and secondary outcomes for patients included in the present analysis DisCoVeRy trial, according to disease severity at baseline. Analyses were stratified on the disease severity at baseline (moderate: 7-point ordinal scale 3 or 4; severe: 7-point ordinal scale 5 or 6), and adjusted effect measures are reported in the table. NP, Nasopharyngeal; LRT, Lower respiratory tract; OR, Odds-ratio; HR, Hazard ratio; LSMD, least-square mean difference.

**Figure 1.**
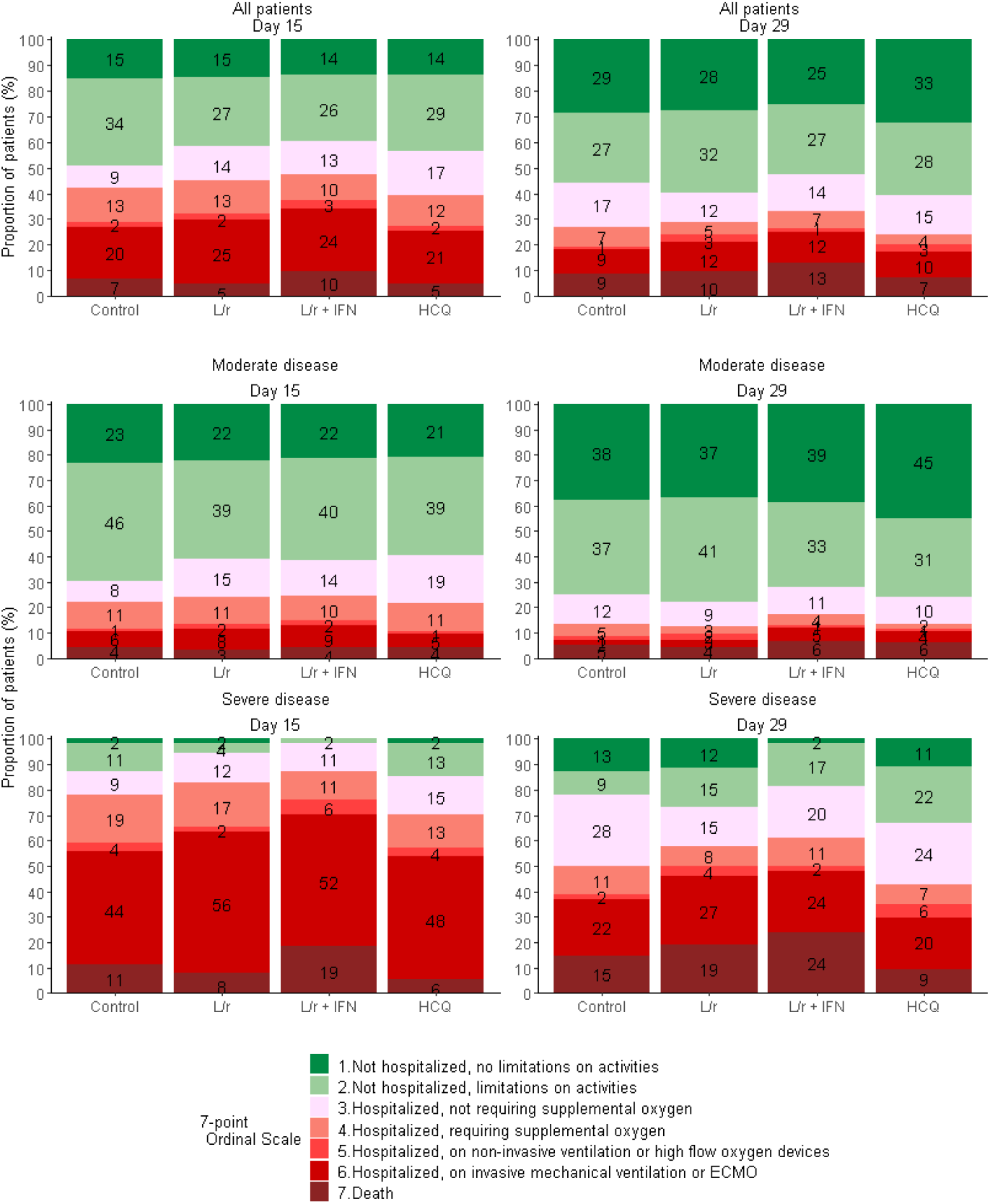
Clinical status, as measured by the 7-point ordinal scale, at day 15 and day 29 of patients from the intention-to-treat population of the DisCoVeRy trial, according to treatment arm and disease severity at baseline. Reported numbers refer to the proportion of patients with the corresponding level in each group. L/r, Lopinavir/ritonavir; L/r + IFN, Lopinavir/ritonavir + interferon ß-1a; HCQ, Hydroxychloroquine.

### Secondary endpoints

There was no significant difference between any of the treatment and control arms on the 7-point ordinal scale at day 29 (Figure 1 and Table 2). The time to improvement of 2 categories of the same scale or hospital discharge within day 29 was significantly longer in lopinavir/ritonavir-containing arms than in the control arm: lopinavir/ritonavir *versus* control, HR=0.70 (95%CI 0.54-0.92, P=0.01 and lopinavir/ritonavir plus IFN-ß-1a *versus* control, HR=0.68 (95%CI 0.52-0.89, P=0.008). The time to NEWS ≤2 or hospital discharge within 29 days was significantly longer in the lopinavir/ritonavir plus IFN-ß-1a arm than in the control arm (HR=0.72, 95%CI 0.54-0.96, P=0.02), as was the time to hospital discharge within day 29 (HR=0.72, 95%CI 0.54-0.97, P=0.03). Participants assigned to the lopinavir/ritonavir plus IFN-ß-1a arm exhibited a higher risk of 3-month mortality than participants assigned to the control arm: aOR 2.11 (95%CI 1.01;4.39, P=0.04), while no significant effect was observed in the lopinavir/ritonavir arm (aOR 1.41, 95%CI 0.65;3.06, P=0.39) nor in the hydroxychloroquine arm (aOR 1.17, 95%CI 0.53;2.58, P=0.70).

No other significant difference was observed for other secondary outcomes (Table 2 and Supplementary Figures S2-S4).

### Virological endpoints

The slope of the decrease of the viral loads in NPS over time was not significantly affected by any of the investigational treatments (Figure 2). No significant difference in the proportion of participants with detectable viral loads at each sampling time was observed in the NPS nor in the LRT specimens (Supplementary Table S2 and S3).

**Figure 2.**
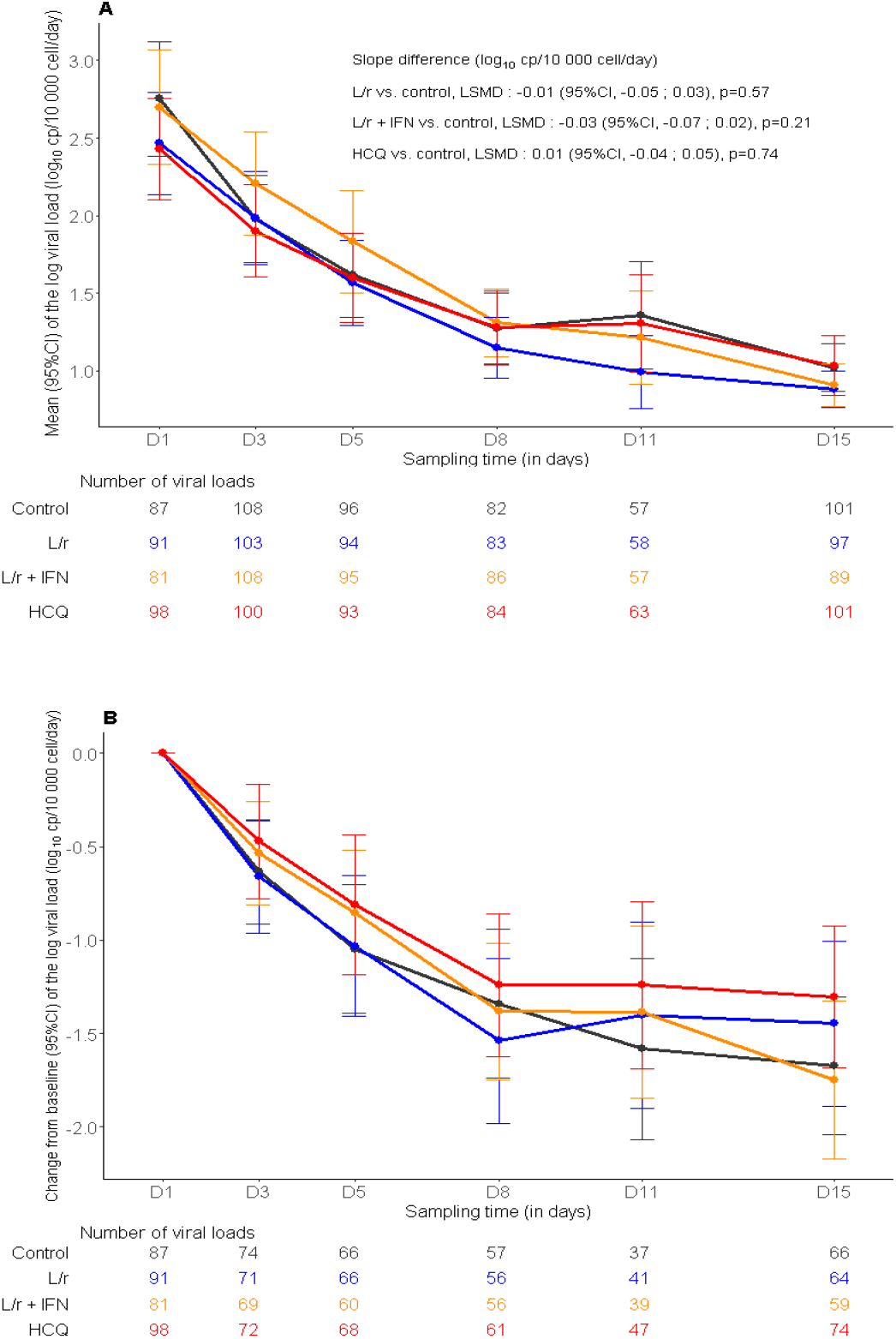
Evolution of the normalized SARS-CoV-2 viral load in nasopharyngeal swabs between baseline and day 15 in the intention-to-treat population of the DisCoVeRy trial: means (95%CI) of the log viral loads (panel A), mean changes from baseline (95%CI) of the log viral loads (panel B). L/r, Lopinavir/ritonavir (blue line); L/r + IFN, Lopinavir/ritonavir + interferon ß-1a (yellow line); HCQ, Hydroxychloroquine (red line); control (black line). LSMD, least-square mean difference; 95%CI, 95% confidence interval.

### Trough concentrations of experimental treatments

At day 3, median trough plasma concentrations of lopinavir were 20 328 ng/mL (IQR, 13 251; 26 980) and 20 926 ng/mL (16 510; 25 930) and of ritonavir were 536 ng/mL (312; 1 028) and 609 ng/mL (388; 1 164) in the lopinavir/ritonavir and in the lopinavir/ritonavir plus IFN-β-1a, respectively (Supplementary Table S4). Median trough plasma concentrations of hydroxychloroquine were 126 ng/mL (67; 276).

### Safety

The safety analysis included 589 participants (control, n=149; lopinavir/ritonavir, n=147; lopinavir/ritonavir plus IFN-β-1a, n=145; hydroxychloroquine, n=148). Safety outcomes are presented in Table 3. Among 2524 reported AEs, 570 were graded 3 or 4 in 243 patients and mostly reported in lopinavir/ritonavir-containing arms (Table 3).

**Table 3.** Summary of adverse events according treatment group in the modified intention to treat population. In the “Overall” column, numbers refer to number of events and number of patients. In other columns, number refer to number of patients (%). Some patients had more than a single SAE. Analyses were performed on the modified Intention-to-treat population. SAE, Serious Adverse Event. P-value refer to Fisher exact test. * According to the investigator’ judgement. Among participants with the occurrence of the SAE related to the experimental treatment, 16 (44.4%) in the lopinavir/ritonavir arm, 36 (65.4%) in the lopinavir/ritonavir plus INF-β-1a arm and 13 (44.8%) in the hydroxychloroquine arm discontinued the experimental treatment. ** Including renal failure in 30 patients, hepatic disorders in 25 patients and electrocardiogram abnormalities in 9 patients. IFN treatment was completed in all patients from the Lopinavir/ritonavir + Interferon ß-1a arm. *** Excluding acute renal failures defined based on the RIFLE classification.

|  | **Overall**<br>**(N=589)** | **Control**<br>**(N=149)** | **Lopinavir/ritonavir**<br>**(L/r)**<br>**(N=147)** | **Lopinavir/ritonavir +**<br>**Interferon β-1a**<br>**(L/r + IFN)**<br>**(N=145)** | **Hydroxychloroquine**<br>**(HCQ)**<br>**(N=148)** | **L/r**<br>**vs. control**<br>**P-value** | **L/r + IFN**<br>**vs. control**<br>**P-value** | **HCQ**<br>**vs. control**<br>**P-value** |
| --- | --- | --- | --- | --- | --- | --- | --- | --- |
|  | **no. events /**<br>**no. patients** | **no.**<br>**patients**<br>**(%)** | **no. patients (%)** | **no. patients (%)** | **no. patients (%)** |  |  |  |
| **Any adverse events** | 2524 / 474 | 113<br>(75.8%) | 125 (85.0%) | 122 (84.1%) | 114 (77.0%) | 0.047 | 0.076 | 0.81 |
| **Any grade 3 or 4 adverse events** | 570 / 243 | 59 (39.6%) | 62 (42.2%) | 65 (44.8%) | 57 (38.5%) | 0.65 | 0.36 | 0.85 |
| **Any serious adverse events** | 856 / 282 | 58 (38.9%) | 76 (51.7%) | 80 (55.2%) | 68 (45.9%) | 0.027 | 0.0053 | 0.22 |
| **Any serious adverse event related**<br>**to the experimental treatment\*** | - | - | 36 (24.5%) | 55 (37.9%) | 29 (19.6%) | - | - | - |
| **Death related to the experimental**<br>**treatment\*** | - | - | 2 (1.4%) | 3 (2.0%) | 0 (0%) | - | - | - |
| **Premature suspension or** | 77 (13.1%) | - | 18 (12.2%) | 42 (29.0%) | 17 (11.5%) | - | - | - |
| **discontinuation of the experimental treatment\*\*** |  |  |  |  |  |  |  |  |
| **Most relevant SAEs** |  |  |  |  |  |  |  |  |
| - Acute respiratory failure | 78 / 65 | 22 (15%) | 18 (12%) | 13 (9%) | 12 (8%) |  |  |  |
| - Acute Respiratory Distress syndrome | 74 / 58 | 15 (10%) | 11 (7%) | 17 (12%) | 15 (10%) |  |  |  |
| - Acute kidney injury*** | 64 / 52 | 9 (6%) | 17 (12%) | 12 (8%) | 14 (9%) |  |  |  |
| - Acute renal failure based on the RIFLE classification | 23 / 16 | 2 (1%) | 3 (2%) | 15 (10%) | 3 (2%) |  |  |  |
| - Arrhythmia | 58 / 38 | 4 (3%) | 8 (5%) | 13 (9%) | 13 (9%) |  |  |  |
| - Pulmonary embolism | 34 / 29 | 7 (5%) | 11 (7%) | 5 (3%) | 6 (4%) |  |  |  |
| - Transaminases increased | 35 / 24 | 2 (1%) | 5 (3%) | 11 (8%) | 6 (4%) |  |  |  |
| - Sepsis | 32 / 25 | 3 (2%) | 9 (6%) | 7 (5%) | 6 (4%) |  |  |  |
| - Cholestasis | 9 / 6 | 0 (0%) | 2 (1%) | 3 (2%) | 1 (1%) |  |  |  |

A total of 856 SAEs were reported in 282 participants; 189 (22.1%) were related to the investigational drug according to investigator’s judgment (lopinavir/ritonavir arm, n=54 lopinavir/ritonavir plus IFN-β-1a arm, n=89; hydroxychloroquine arm, n=46). A significantly greater number of patients experienced at least one SAE in the lopinavir/ritonavir-containing arms than in the control arm (Table 3). The most frequently reported SAEs were acute respiratory failure (n=78, 13.2%), acute kidney injury (n=64, 10.8%), acute respiratory distress syndrome (n=74, 12.5%), arrhythmia (n=58, 9.4%), pulmonary embolism (n=34, 55.7%), and sepsis including those related to super-infections (n=32, 5.4%). Twelve percent (n=103) of participants developed at least one kidney-related SAE. Among them, 23 had acute renal failure upon admission, and 78 were critically-ill ventilated patients with acute kidney injury. Among 64 fatal SAEs, 32 had a pulmonary origin, and 32 had a non-pulmonary origin. Three non-pulmonary-related deaths were linked to investigational treatments by investigators, all in the - lopinavir/ritonavir plus IFN-ß-1a arm.

## Discussion

We report here the results of the DisCoVeRy clinical trial, evaluating lopinavir/ritonavir with or without IFN-ß-1a, or hydroxychloroquine in comparison with control for the treatment of inpatients with COVID-19. Participants had mostly moderate disease (63.9%) covering a large spectrum of clinical presentations. Inclusions were prematurely stopped for futility, so that the number of included patients is lower than the estimated sample size. Consistently with Solidarity results, investigational treatments failed to improve the clinical course of COVID-19. No effect on SARS-CoV-2 clearance was observed, using a reproducible normalized method. Furthermore, significantly more SAEs were reported in the lopinavir/ritonavir-containing arms than in the control arm.

Two randomized trials conducted in hospitalized COVID-19 patients found no benefit of lopinavir/ritonavir in terms of 28-day mortality or of progression to mechanical ventilation or death (9,21). No added benefit was observed using IFN-ß-1a, as the median time to randomization of 9 days may have been too long to allow an immune-mediated boosting effect on viral clearance. We observed plasma overexposure of lopinavir relative to target concentrations obtained in HIV-infected patients, possibly responsible for the higher rate of SAEs and more acute kidney injury than controls. The SARS-CoV-2-induced inflammatory burden may have reduced Cytochrome P450 activity and modified plasma α-1-acid glycoprotein levels, an acute phase protein which binds protease inhibitors (22,23). Reported *in-vitro* half maximal inhibitory concentration (EC50) for SARS-CoV2 is 16,400 ng/mL (24) (while the EC50 for HIV is 70 ng/mL (25)), an over 200-fold difference, suggesting that significantly higher concentrations of lopinavir are needed to enhance SARS-CoV-2 clearance. A recent physiologically-based pharmacokinetic model suggested that standard regimens of lopinavir/ritonavir are not sufficient to achieve efficacy through unbound lung concentrations (26). In our study, trough lopinavir plasma concentrations at day 3 were more than 2-fold higher than expected with the standard dose (27), but were below the EC50 of SARS-CoV2 in 25% of participants.

Several larger-scale randomized controlled trials conducted in hospitalized COVID-19 patients failed to demonstrate the clinical efficacy of hydroxychloroquine (28,29). Our results are in line with these conclusions. We report that hydroxychloroquine does not accelerate SARS-CoV-2 clearance, consistent with preclinical data (30). Based on *in vitro* EC50 against SARS-CoV-2 (242 ng/mL), the target plasma concentration was reached in only 25% of participants at day 3, and optimal intrapulmonary exposure might have been only achieved at day 10 (10,17). It could be argued that the dosing regimen administered in the DisCoVeRy trial was insufficient to rapidly reach target concentrations. However, Solidarity and Recovery trials, which both used a doubled hydroxychloroquine dosing regimen, did not bring evidence of clinical benefit either (13,29).

The trial has limitations: the complexity of blinding treatments with different routes of administration and the need to initiate the trial very rapidly led to choose an open-labelled design. The trial did not target patients at the early phase of the disease nor include arms testing anti-inflammatory agents that could be used as part of the SoC in any arm. In addition, the trial was performed in the early phase of the COVID-19 pandemics and the SoC underwent substantial changes over time, adapting to knowledge acquisition, especially regarding the use of corticosteroids in COVID-19.

## Conclusion

In patients admitted to hospital with COVID-19, lopinavir/ritonavir, lopinavir/ritonavir plus IFN-β-1a and hydroxychloroquine were not associated with clinical improvement at day 15 and day 29, nor reduction in viral shedding, and generated significantly more SAEs in lopinavir/ritonavir-containing arms. These findings do not support the use of these investigational treatments for patients hospitalized with COVID-19.

## Data Availability

All data produced in the present study are available upon reasonable request to the authors

## Authors contribution

Writing – Original Draft: FA, CB; Writing – Review & Editing: NPS, JP, MBD, DB, ADi, MH, MPL, GP, DC, YY, FM; Conceptualization: FA, NPS, JP, MBD, GP, BL, DC, YY, FM; Investigation: FA, NPS, JP, MBD, Adi, NM, FXL, FR, FG, AK, SJ, JR, SN, FD, RCJ, KB, JCN, VT, AC, CDu, JC, SL, JM, RG, BM, EF, VP, SG, OL, KL, JPL, AM, GMB, LB, ÉBN, AGB, OE, LP, FW, JCR, JR, TS, MH, CA, MPL, GP; Methodology: FA, NPS, JP, MBD, DC, CB, FM; Data curation: ADi, ADe, NM, ADu, TA; Formal Analysis: DB, ADu, DC, CB, FM

Project Administration: FA, CD, FM; Funding Acquisition: FA, CDe, JS, DC, YY, FM.

## References

1. Zhu N, Zhang D, Wang W, Li X, Yang B, Song J, et al. A Novel Coronavirus from Patients with Pneumonia in China, 2019. The New England journal of medicine. 2020;382(8):727□33.

2. Zhou F, Yu T, Du R, Fan G, Liu Y, Liu Z, et al. Clinical course and risk factors for mortality of adult inpatients with COVID-19 in Wuhan, China: a retrospective cohort study. Lancet. 2020;395(10229):1054□62.

3. Chu CM, Cheng VC, Hung IF, Wong MM, Chan KH, Chan KS, et al. Role of lopinavir/ritonavir in the treatment of SARS: initial virological and clinical findings. Thorax. 2004;59(3):252□6.

4. de Wilde AH, Jochmans D, Posthuma CC, Zevenhoven-Dobbe JC, van Nieuwkoop S, Bestebroer TM, et al. Screening of an FDA-approved compound library identifies four small-molecule inhibitors of Middle East respiratory syndrome coronavirus replication in cell culture. Antimicrobial agents and chemotherapy. 2014;58(8):4875□84.

5. Sallard E, Lescure FX, Yazdanpanah Y, Mentre F, Peiffer-Smadja N. Type 1 interferons as a potential treatment against COVID-19. Antiviral research. 2020;178:104791.

6. Lokugamage KG, Hage A, de Vries M, Valero-Jimenez AM, Schindewolf C, Dittmann M, et al. Type I Interferon Susceptibility Distinguishes SARS-CoV-2 from SARS-CoV. J Virol. 9 nov 2020;94(23):e01410–20.

7. Clementi N, Ferrarese R, Criscuolo E, Diotti RA, Castelli M, Scagnolari C, et al. Interferon-beta-1a Inhibition of Severe Acute Respiratory Syndrome-Coronavirus 2 In Vitro When Administered After Virus Infection. The Journal of infectious diseases. 2020;222(5):722□5.

8. Liu J, Cao R, Xu M, Wang X, Zhang H, Hu H, et al. Hydroxychloroquine, a less toxic derivative of chloroquine, is effective in inhibiting SARS-CoV-2 infection in vitro. Cell discovery. 2020;6:16.

9. Cao B, Wang Y, Wen D, Liu W, Wang J, Fan G, et al. A Trial of Lopinavir-Ritonavir in Adults Hospitalized with Severe Covid-19. N Engl J Med. 7 mai 2020;382(19):1787□99.

10. Yao X, Ye F, Zhang M, Cui C, Huang B, Niu P, et al. In Vitro Antiviral Activity and Projection of Optimized Dosing Design of Hydroxychloroquine for the Treatment of Severe Acute Respiratory Syndrome Coronavirus 2 (SARS-CoV-2). Clinical infectious diseases[: an official publication of the Infectious Diseases Society of America. 2020;71(15):732□9.

11. Warren TK, Jordan R, Lo MK, Ray AS, Mackman RL, Soloveva V, et al. Therapeutic efficacy of the small molecule GS-5734 against Ebola virus in rhesus monkeys. Nature. 2016;531(7594):381□5.

12. Ader F, Discovery French Trial Management T. Protocol for the DisCoVeRy trial: multicentre, adaptive, randomised trial of the safety and efficacy of treatments for COVID-19 in hospitalised adults. BMJ open. 2020;10(9):e041437.

13. WHO Solidarity Trial Consortium, Pan H, Peto R, Henao-Restrepo A-M, Preziosi M-P, Sathiyamoorthy V, et al. Repurposed Antiviral Drugs for Covid-19 - Interim WHO Solidarity Trial Results. N Engl J Med. 11 févr 2021;384(6):497□511.

14. Ader F, Peiffer-Smadja N, Poissy J, Bouscambert-Duchamp M, Belhadi D, Diallo A, et al. An open-label randomized controlled trial of the effect of lopinavir/ritonavir, lopinavir/ritonavir plus IFN-β-1a and hydroxychloroquine in hospitalized patients with COVID-19. Clin Microbiol Infect. 26 mai 2021;S1198-743X(21)00259-7.

15. Ader F, Bouscambert-Duchamp M, Hites M, Peiffer-Smadja N, Poissy J, Belhadi D, et al. Remdesivir plus standard of care versus standard of care alone for the treatment of patients admitted to hospital with COVID-19 (DisCoVeRy): a phase 3, randomised, controlled, open-label trial. The Lancet Infectious Diseases. févr 2022;22(2):209□21.

16. Arabi YM, Alothman A, Balkhy HH, Al-Dawood A, AlJohani S, Al Harbi S, et al. Treatment of Middle East Respiratory Syndrome with a combination of lopinavir-ritonavir and interferon-beta1b (MIRACLE trial): study protocol for a randomized controlled trial. Trials. 2018;19(1):81.

17. Le MP, Peiffer-Smadja N, Guedj J, Neant N, Mentre F, Ader F, et al. Rationale of a loading dose initiation for hydroxychloroquine treatment in COVID-19 infection in the DisCoVeRy trial. The Journal of antimicrobial chemotherapy. 2020;75(9):2376□80.

18. Etievant S, Bal A, Escuret V, Brengel-Pesce K, Bouscambert M, Cheynet V, et al. Performance Assessment of SARS-CoV-2 PCR Assays Developed by WHO Referral Laboratories. J Clin Med. 16 juin 2020;9(6):E1871.

19. Jung BH, Rezk NL, Bridges AS, Corbett AH, Kashuba ADM. Simultaneous determination of 17 antiretroviral drugs in human plasma for quantitative analysis with liquid chromatography–tandem mass spectrometry. Biomed Chromatogr. oct 2007;21(10):1095□104.

20. Chhonker YS, Sleightholm RL, Li J, Oupický D, Murry DJ. Simultaneous quantitation of hydroxychloroquine and its metabolites in mouse blood and tissues using LC–ESI–MS/MS: An application for pharmacokinetic studies. Journal of Chromatography B. janv 2018;1072:320□7.

21. Horby PW, Mafham M, Bell JL, Linsell L, Staplin N, Emberson J, et al. Lopinavir–ritonavir in patients admitted to hospital with COVID-19 (RECOVERY): a randomised, controlled, open-label, platform trial. The Lancet. oct 2020;396(10259):1345□52.

22. Marzolini C, Stader F, Stoeckle M, Franzeck F, Egli A, Bassetti S, et al. Effect of Systemic Inflammatory Response to SARS-CoV-2 on Lopinavir and Hydroxychloroquine Plasma Concentrations. Antimicrob Agents Chemother. 20 août 2020;64(9):e01177–20.

23. Ofotokun I, Lennox JL, Eaton ME, Ritchie JC, Easley KA, Masalovich SE, et al. Immune activation mediated change in alpha-1-acid glycoprotein: impact on total and free lopinavir plasma exposure. J Clin Pharmacol. nov 2011;51(11):1539□48.

24. Choy KT, Wong AY, Kaewpreedee P, Sia SF, Chen D, Hui KPY, et al. Remdesivir, lopinavir, emetine, and homoharringtonine inhibit SARS-CoV-2 replication in vitro. Antiviral research. 2020;178:104786.

25. Croxtall JD, Perry CM. Lopinavir/Ritonavir: a review of its use in the management of HIV-1 infection. Drugs. 2010;70(14):1885□915.

26. Thakur A, Tan SPF, Chan JCY. Physiologically-Based Pharmacokinetic Modeling to Predict the Clinical Efficacy of the Coadministration of Lopinavir and Ritonavir against SARS-CoV-2. Clinical pharmacology and therapeutics. 2020;108(6):1176□84.

27. Kaletra. Summary of Product Characteristics. [Internet]. [accessed Feb 15, 2022]. Available at: https://www.ema.europa.eu/en/documents/product-information/kaletra-epar-product-information_en.pdf

28. Cavalcanti AB, Zampieri FG, Rosa RG, Azevedo LCP, Veiga VC, Avezum A, et al. Hydroxychloroquine with or without Azithromycin in Mild-to-Moderate Covid-19. The New England journal of medicine. 2020;383(21):2041□52.

29. RECOVERY Collaborative Group, Horby P, Mafham M, Linsell L, Bell JL, Staplin N, et al. Effect of Hydroxychloroquine in Hospitalized Patients with Covid-19. N Engl J Med. 19 nov 2020;383(21):2030□40.

30. Maisonnasse P, Guedj J, Contreras V, Behillil S, Solas C, Marlin R, et al. Hydroxychloroquine use against SARS-CoV-2 infection in non-human primates. Nature. sept 2020;585(7826):584□7.

