## Supplemental material for "An open-label randomized, controlled trial of the effect of lopinavir/ritonavir, lopinavir/ritonavir plus IFN-β-1a and hydroxychloroquine in hospitalized patients with COVID-19 – Final results from the DisCoVeRy trial"

**Supplementary Appendix**

**Supplementary Methods**

***Selection criteria***

We enrolled adult patients who were hospitalized with PCR-confirmed COVID-19, as determined by a positive SARS-CoV-2 RT-PCR on any biological sample.

The full list of inclusion and exclusion criteria was the following.

Inclusion criteria:

- Adult ≥18 years of age at time of enrolment;
- laboratory-confirmed SARS-CoV-2 infection as determined by PCR, or other commercial or public health assay in any specimen < 72 hours prior to randomization;
- Hospitalized patients with illness of any duration, and at least one of the following:
  - Clinical assessment (evidence of rales/crackles on physical examination) AND SpO2 ≤ 94% on room air, OR
  - Acute respiratory failure requiring supplemental oxygen, high flow oxygen devices, non-invasive ventilation, and/or mechanical ventilation;
- Women of childbearing potential must agree to use contraception for the duration of the study.

Exclusion criteria:

- Refusal to participate expressed by patient or legally authorized representative if they are present;
- Spontaneous blood alanine transferase (ALT)/AST levels > 5 times the upper limit of normal;
- Stage 4 severe chronic kidney disease or requiring dialysis (i.e. eGFR < 30 mL/min);
- Pregnancy or breast-feeding;
- Anticipated transfer to another hospital, which is not a study site within 72 hours;
- Patients previously treated with one of the antivirals evaluated in the trial (i.e. remdesivir, interferon ß-1a, lopinavir/ritonavir, hydroxychloroquine) in the past 29 days;
- Contraindication to any study medication including allergy;
- Use of medications that are contraindicated with lopinavir/ritonavir i.e. drugs whose metabolism is highly dependent on the isoform CYP3A with narrow therapeutic range (e.g. amiodarone, colchicine, simvastatine);
- Use of medications that are contraindicated with hydroxychloroquine: citalopram, escitalopram, hydroxyzine, domperidone, pipéraquine;
- Human immunodeficiency virus infection under highly active antiretroviral therapy (HAART);
- History of severe depression or attempted suicide or current suicidal ideation;
- Corrected QT interval superior to 500 milliseconds (as calculated with the Fridericia formula).

***Interim analyses***

For efficacy and futility, the statistical analysis was performed on the primary outcome measure, and was based on the Haybittle-Peto rule ^1^^,^^2^. As an add-on trial, DisCoVeRy periodically transferred data to the WHO Solidarity trial (in-hospital mortality, time to hospital discharge, time to mechanical ventilation), whose DSMB examined all Solidarity trial data.

On May 25^th^ 2020, following a safety warning on hydroxychloroquine use ^3^, enrollment in the hydroxychloroquine arm was suspended at the request of the French Agency of drug Security (Agence Nationale de Sécurité du Médicament). On June 13^th^, based on the interim analysis of the Solidarity data, the Solidarity and DisCoVeRy trial DSMBs recommended to definitely stop the hydroxychloroquine arm due to futility. This decision was endorsed by the DisCoVeRy steering committee on June 17^th^. The Solidarity DSMB advised to stop the lopinavir/ritonavir arm due to futility on June, 23^th^. Thereafter, the DisCoVeRy DSMB further advised to stop both the lopinavir/ritonavir-containing arms due to additional safety concerns on June, 25^th^. This decision was endorsed by the DisCoVeRy steering committee on June 27^th^ with subsequent interruption on June, 29^th^.

***Statistical analyses***

Statistical analyses compared each of the three stopped investigational treatment arms to the control arm. The intention-to-treat population included all randomized participants for whom a valid consent form was obtained. The modified intention-to-treat population included participants from the intention-to-treat population who received at least one dose of the treatment allocated by randomization.

Efficacy analyses were performed on the intention-to-treat population; handling of missing data is described in Supplementary Appendix. Safety analyses were performed on modified intention-to-treat population. Analyses were stratified by baseline severity but not by region of inclusion due to a low number of inclusions in some regions; all tests were two-sided with a type-I error of 0.05, without correction for multiplicity testing.

For the 7-point ordinal scale, data were analyzed using a proportional odds model. Time-to-event data were analyzed using a Cox proportional hazard model. An analysis of covariance was performed for the comparison of oxygenation- and ventilator-free days between groups; 29-day mortality and the number of participants with detectable SARS-CoV-2 in the respiratory tract specimens at each time point were analyzed using a Cochran-Mantel-Haenszel test. Log_10_ normalized SARS-CoV-2 load kinetics was analyzed using a mixed-effects linear model with test of treatment effect on slopes. For safety endpoints, non-prespecified statistical comparisons of the proportions of patients with any i) adverse event, ii) grade 3 or 4 adverse event, or iii) serious adverse event between each investigational treatment arm versus control were performed using the Fisher exact test.

***Handling of missing data***

For the 7-point ordinal scale, missing data were imputed using the last observation carried forward method, excepted in the case of known death or hospital discharge, in which case the ordinal scale was imputed to the value of 7 (death) or 2 (not hospitalized, limitation of activities), respectively. For NEWS, oxygenation and mechanical ventilation outcomes, missing data were treated using the last observation carried forward method, except in the case of preceding death, in which case patients were imputed to the worst value of the NEWS, or considered to have oxygen or require mechanical ventilation. For time-to-event analyses, patients were censored at day 29, at their date of loss of follow-up, or of study withdrawal, whichever occurred first. As time to event analyses were performed only for positive events (time to an improvement of 2 categories as measured on the 7-point ordinal scale or hospital discharge until day 29, time to National Early Warning Score 2 (NEWS2) ≤2 or hospital discharge until day 29, and time to hospital discharge until day 29), patients who died before the day 29 visit were considered as patients at risk in the survival analysis and censored at day 29. Missing SARS-CoV-2 viral loads were not imputed. For the analysis of viral load by mixed models, undetectable viral load values (i.e. values < 1 log10 copies/10 000 cells) were imputed to half the LoD hence 0.7 log10 copies/10 000 cells. In case of several consecutive undetectable values, only the first one was replaced and the subsequent discarded (until the next detectable value, if values were available afterwards).

**Supplementary Figures**

**Supplementary Figure S1. Enrollment and randomization of patients in the present analysis of the DisCoVeRy trial.**

Patients from all groups received the standard of care, in addition to the treatment allocated by randomization.

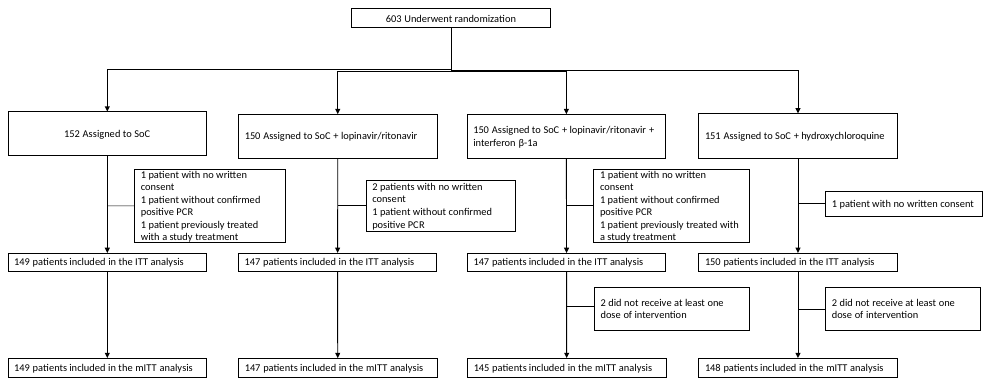

**Supplementary Figure S2. Time to improvement of at least 2 categories or the 7-point ordinal scale or hospital discharge between baseline and day 29 in the intention-to-treat population of the DisCoVeRy trial, according to disease severity at baseline in all participants (panel A), in participants with moderate disease at baseline (panel B) and in participants with severe disease at baseline (panel C).**

L/r, Lopinavir/ritonavir; L/r + IFN, Lopinavir/ritonavir + interferon ß-1a; HCQ, Hydroxychloroquine; HR, hazard ratio.

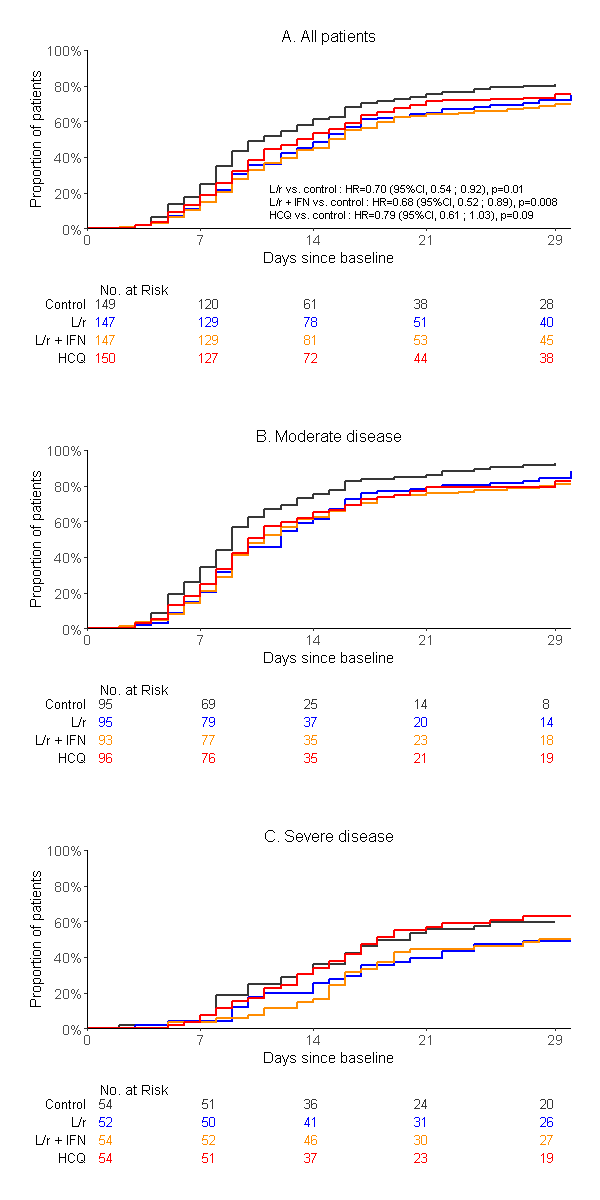

**Supplementary Figure S3. Time to National Early Warning Score ≤2 or hospital discharge between baseline and day 29 in the intention-to-treat population of the DisCoVeRy trial, according to disease severity at baseline in all participants (panel A), in participants with moderate disease at baseline (panel B) and in participants with severe disease at baseline (panel C).**

L/r, Lopinavir/ritonavir; L/r + IFN, Lopinavir/ritonavir + interferon ß-1a; HCQ, Hydroxychloroquine; HR, hazard ratio.

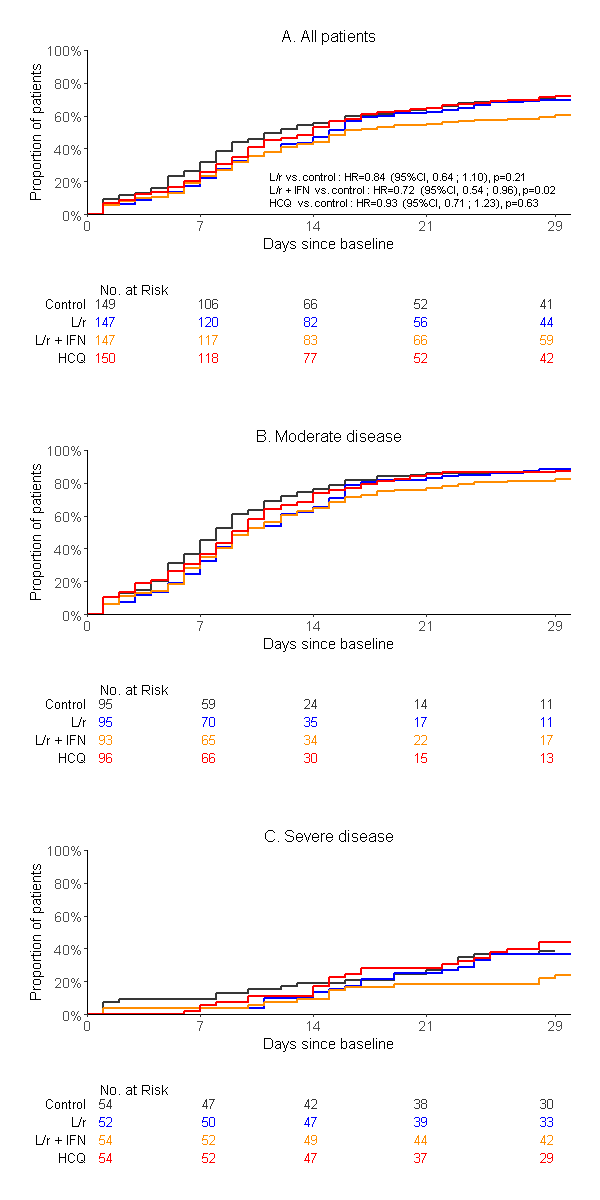

**Supplementary Figure S4. Time to hospital discharge within day 29 between baseline and day 29 in the intention-to-treat population of the DisCoVeRy trial, according to disease severity at baseline in all participants (panel A), in participants with moderate disease at baseline (panel B) and in participants with severe disease at baseline (panel C).**

L/r, Lopinavir/ritonavir; L/r + IFN, Lopinavir/ritonavir + interferon ß-1a; HCQ, Hydroxychloroquine; HR, hazard ratio.

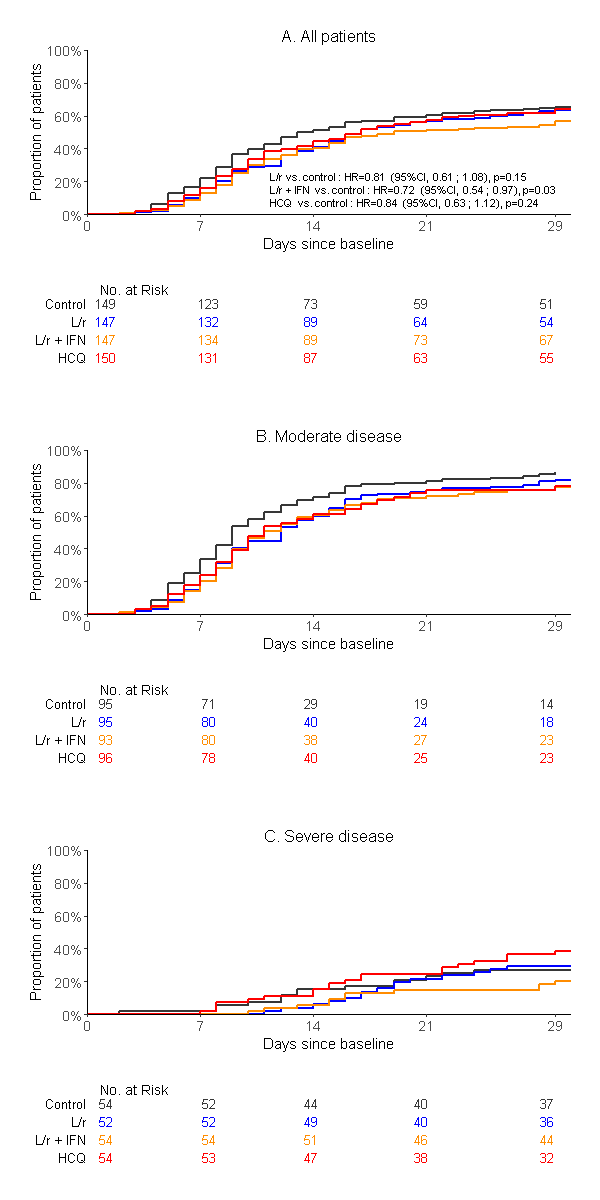

**Supplementary Tables**

**Supplementary Table S1. Treatments received during the study course in the patients included in the intention-to-treat population of the DisCoVeRy trial.**

| **Treatment — no. (%)** | **Overall**  **(N=593)** | **Control**  **(N=149)** | **Lopinavir/ritonavir**  **(L/r)**  **(N=147)** | **Lopinavir/ritonavir + interferon ß-1a**  **(L/r + IFN)**  **(N=147)** | **Hydroxychloroquine**  **(HCQ)**  **(N=150)** |
| --- | --- | --- | --- | --- | --- |
| **Corticosteroids** | 176 (29.9%) | 41 (27.7%) | 45 (30.8%) | 48 (33.1%) | 42 (28.0%) |
| - Dexamethasone | 45 (7.6%) | 9 (6.1%) | 11 (7.5%) | 13 (9.0%) | 12 (8.0%) |
| - Hydrocortisone | 30 (5.1%) | 7 (4.7%) | 9 (6.2%) | 8 (5.5%) | 6 (4.0%) |
| - Methyprednisolone | 69 (11.7%) | 16 (10.8%) | 19 (13.0%) | 19 (13.1%) | 15 (10.0%) |
| **Interleukin-6 inhibitors** | 1 (0.2%) | 1 (0.7%) | 0 (0.0%) | 0 (0.0%) | 0 (0.0%) |
| - Tocilizumab | 1 (0.2%) | 1 (0.7%) | 0 (0.0%) | 0 (0.0%) | 0 (0.0%) |
| - Sarilumab | 0 (0.0%) | 0 (0.0%) | 0 (0.0%) | 0 (0.0%) | 0 (0.0%) |
| **Interleukin-1 inhibitors** | 6 (1.0%) | 1 (0.7%) | 1 (0.7%) | 2 (1.4%) | 2 (1.3%) |
| **Antibiotics** | 469 (79.3%) | 117 (78.5%) | 120 (81.6%) | 117 (80.7%) | 115 (76.8%) |
| - Azithromycine | 18 (3.05%) | 4 (2.7%) | 7 (4.7%) | 3 (2.1%) | 4 (2.7%) |
| **Anticoagulants** | 557 (94.6%) | 135 (91.2%) | 138 (94.5%) | 137 (94.5%) | 147 (98.0%) |
| **Parenteral or enteral nutrition** | 234 (39.7%) | 56 (37.8%) | 63 (43.1%) | 64 (44.1%) | 51 (34.0%) |
| **Vasopressors** | 222 (37.7%) | 52 (35.1%) | 56 (38.7%) | 63 (43.4%) | 51 (34.0%) |
| **Extra-renal replacement/hemofiltration** | 56 (9.5%) | 8 (5.4%) | 18 (12.3%) | 22 (15.2%) | 8 (5.3%) |
| **Neuromuscular blocking agents** | 199 (33.8%) | 48 (32.4%) | 45 (30.8%) | 59 (40.7%) | 47 (31.3%) |
| **Inhaled nitric oxide** | 44 (7.5%) | 8 (5.4%) | 10 (6.8%) | 17 (11.7%) | 9 (6.0%) |
| **Prone positioning** | 189 (32.1%) | 37 (25.0%) | 44 (30.1%) | 66 (45.5%) | 42 (28.0%) |

**Supplementary Table S2. Proportion of patients with detectable viral loads in the nasopharyngeal swabs at each sampling time, in the intention-to-treat population of the DisCoVeRy trial.**

NPS, Nasopharyngeal swab.

|  | **Overall**  **(N=593)** | | **Control**  **(N=149)** | | **Lopinavir/ritonavir**  **(L/r)**  **(N=147)** | | **Lopinavir/ritonavir + interferon ß-1a**  **(L/r + IFN)**  **(N=147)** | | **Hydroxychloroquine**  **(HCQ)**  **(N=150)** | | **L/r**  **vs. control**  **Effect measure**  **(95%CI)** | **L/r + IFN**  **vs. control**  **Effect measure**  **(95%CI)** | **HCQ**  **vs. control**  **Effect measure**  **(95%CI)** |
| --- | --- | --- | --- | --- | --- | --- | --- | --- | --- | --- | --- | --- | --- |
|  | **Moderate (N=379)** | **Severe (N=214)** | **Moderate (N=95)** | **Severe (N=54)** | **Moderate (N=95)** | **Severe (N=52)** | **Moderate (N=93)** | **Severe (N=54)** | **Moderate (N=96)** | **Severe (N=54)** |  |  |  |
| **Detectable viral load in NPS at day 3**, n/N (%) | 154/277 (55.6%) | 83/131 (63.4%) | 35/68 (51.5%) | 25/34 (73.5%) | 38/69 (55.1%) | 21/32 (65.6%) | 43/72 (59.7%) | 20/33 (60.6%) | 38/68 (55.9%) | 17/32 (53.1%) | OR=0.99 (0.57 to 1.75)  [P=0.98] | OR=1.06 (0.61 to 1.85)  [P=0.84] | OR=0.86 (0.49 to 1.50)  [P=0.60] |
| **Detectable viral load in NPS at day 5**, n/N (%) | 107/257 (41.6%) | 59/116 (50.9%) | 22/64 (34.4%) | 18/31 (58.1%) | 25/65 (38.5%) | 14/29 (48.3%) | 33/65 (50.8%) | 15/27 (55.6%) | 27/63 (42.9%) | 12/29 (41.4%) | OR=0.99 (0.55 to 1.77)  [P=0.96] | OR=1.54 (0.86 to 2.75)  [P=0.15] | OR=1.02 (0.57 to 1.82)  [P=0.95] |
| **Detectable viral load in NPS at day 8**, n/N (%) | 64/210 (30.5%) | 45/122 (36.9%) | 16/51 (31.4%) | 10/30 (33.3%) | 17/53 (32.1%) | 6/30 (20.0%) | 18/50 (36.0%) | 15/34 (44.1%) | 13/56 (23.2%) | 14/28 (50.0%) | OR=0.81 (0.41 to 1.58)  [P=0.54] | OR=1.36 (0.72 to 2.58)  [P=0.35] | OR=1.03 (0.53 to 1.98)  [P=0.94] |
| **Detectable viral load in NPS at day 11,** n/N (%) | 26/122 (21.3%) | 30/105 (28.6%) | 6/29 (20.7%) | 7/25 (28.0%) | 4/32 (12.5%) | 5/26 (19.2%) | 5/27 (18.5%) | 10/27 (37.0%) | 11/34 (32.4%) | 8/27 (29.6%) | OR=0.58 (0.22 to 1.50)  [P=0.26] | OR=1.19 (0.50 to 2.84)  [P=0.70] | OR=1.43 (0.63 to 3.26)  [P=0.40] |
| **Detectable viral load in NPS at day 15**, n/N (%) | 41/272 (15.1%) | 24/116 (20.7%) | 12/69 (17.4%) | 8/32 (25.0%) | 8/73 (11.0%) | 5/26 (19.2%) | 10/63 (15.9%) | 2/24 (8.3%) | 11/67 (16.4%) | 9/34 (26.5%) | OR=0.63 (0.29 to 1.35)  [P=0.24] | OR=0.65 (0.30 to 1.42)  [P=0.28] | OR=0.99 (0.49 to 1.98)  [P=0.98] |
| **Detectable viral load in NPS at day 29**, n/N (%) | 11/242 (4.5%) | 4/90  (4.4%) | 3/62 (4.8%) | 2/19 (10.5%) | 3/64 (4.7%) | 1/22 (4.5%) | 2/58 (3.4%) | 0/24 (0.0%) | 3/58 (5.2%) | 1/25  (4.0%) | OR=0.73 (0.19 to 2.83)  [P=0.65] | OR=0.39 (0.08 to 2.01)  [P=0.24] | OR=0.75 (0.20 to 2.89)  [P=0.68] |

**Supplementary Table S3. Number of severe patients with detectable normalized viral load in the lower respiratory tract at each sampling time, in the intention-to-treat population of the DisCoVeRy trial.**

|  | **Overall**  **(N=211)** | **Control**  **(N=54)** | **Lopinavir/ritonavir**  **(L/r)**  **(N=51)** | **Lopinavir/ritonavir + interferon**  **(L/r + IFN)**  **(N=54)** | **Hydroxychloroquine (HCQ)**  **(N=52)** |
| --- | --- | --- | --- | --- | --- |
| **Detectable viral load in LRT samples at day 3,** n/N (%) | 33/35 (94.3%) | 9/9 (100.0%) | 9/10 (90.0%) | 10/11 (90.9%) | 5/5 (100.0%) |
| **Detectable viral load in LRT samples at day 5,** n/N (%) | 36/39 (92.3%) | 7/7 (100.0%) | 10/10 (100.0%) | 11/11 (100.0%) | 8/11 (72.7%) |
| **Detectable viral load in LRT samples at day 8,** n/N (%) | 23/30 (76.7%) | 5/7 (71.4%) | 7/9 (77.8%) | 6/7 (85.7%) | 5/7 (71.4%) |
| **Detectable viral load in LRT samples at day 11**, n/N (%) | 16/26 (61.5%) | 5/8 (62.5%) | 5/6 (83.3%) | 4/7 (57.1%) | 2/5 (40.0%) |
| **Detectable viral load in LRT samples at day 15,** n/N (%) | 3/17 (17.6%) | 1/3 (33.3%) | 1/5 (20.0%) | 1/7 (14.3%) | 0/2 (0.0%) |
| **Detectable viral load in LRT samples at day 29**, n/N (%) | 2/9 (22.2%) | 0/2 (0.0%) | 0/3 (0.0%) | 1/2 (50.0%) | 1/2 (50.0%) |

**Supplementary Table S4. Trough plasma concentrations of lopinavir, ritonavir and hydroxychloroquine at day 1 and day 3 in the intention-to-treat population of the DisCoVeRy trial.**

Expected lopinavir concentrations at day 3 were 8100 ng/mL (see ref. 25 in main text).

Data are presented as median (IQR 25-75%).

|  | **Lopinavir/ritonavir**  **arm** | | **Lopinavir/ritonavir + interferon**  **arm** | | **Hydroxychloroquine** |
| --- | --- | --- | --- | --- | --- |
|  | **Lopinavir** | **Ritonavir** | **Lopinavir** | **Ritonavir** |  |
| Trough plasma concentration at day 1 (ng/mL) | 12 880  (8 640; 17 130)  n=86 | 590  (315; 830)  n=71 | 11 061  (7 810; 15 190)  n=74 | 483  (250; 742)  n=63 | 46  (24; 138)  n=62 |
| Trough plasma concentration at day 3 (ng/mL) | 20 328  (13 251; 26 980)  n=70 | 536  (312; 1 028)  n=59 | 20 926  (16 510; 25 930)  n=73 | 609  (388; 1 164)  n=59 | 126  (67; 276)  n=27 |

**The DisCoVeRy Study Group:**

**The French DisCoVeRy Trial Management Team:**

F Ader, Y Yazdanpanah, F Mentre, N Peiffer-Smadja, FX Lescure, J Poissy, L Bouadma, JF Timsit, B Lina, F Morfin-Sherpa, M Bouscambert, A Gaymard, G Peytavin, L Abel, J Guedj, C Andrejak, C Burdet, C Laouenan, D Belhadi, A Dupont, T Alfaiate, B Basli, A Chair, S Laribi, J Level, M Schneider, MC Tellier, A Dechanet, D Costagliola, B Terrier, M Ohana, S Couffin-Cadiergues, H Esperou, C Delmas, J Saillard, C Fougerou, L Moinot, L Wittkop, C Cagnot, S Le Mestre, D Lebrasseur-Longuet, V Petrov-Sanchez, A Diallo, N Mercier, V Icard, B Leveau, S Tubiana, B Hamze, A Gelley, M Noret, E D’Ortenzio, O Puechal, C Semaille.

**The DisCoVeRy Steering Committee (members who are not listed in other groups):**

T Welte, JA Paiva, M Halanova, MP Kieny

**ANRS (France Recherche Nord&Sud SIDA-HIV Hépatites), Paris, France:** E Balssa, C Birkle, S Gibowski, E Landry, A Le Goff, L Moachon, C Moins, L Wadouachi, C Paul, A Levier

**Centre Hospitalier Annecy Genevois, France:** D Bougon

**Centre Hospitalier de Cayenne Andrée Rosemon, Cayenne, France:** F Djossou, L Epelboin

**Centre Hospitalier Universitaire de Nice, France:** J Dellamonica, CH Marquette

**Centre Hospitalier Régional de Metz-Thionville, France:** C Robert

**Centre Hospitalier Régional Universitaire de Nancy, France:** S Gibot

**Centre Hospitalier de Tourcoing, France:** E Senneville, V Jean-Michel

**Centre Hospitalier Universitaire de Amiens, France:** Y Zerbib

**Centre Hospitalier Universitaire de Besançon, France:** C Chirouze

**Centre Hospitalier Universitaire de Bordeaux, France:** A Boyer, C Cazanave, D Gruson, D Malvy

**Centre Hospitalier Universitaire de Dijon, France:** P Andreu, JP Quenot

**Centre Hospitalier Universitaire de Grenoble Alpes, France:** N Terzi

**Centre Hospitalier Universitaire de Lille, France:** K Faure

**Centre Hospitalier Universitaire de Martinique, Fort-de-France, France:** C Chabartier

**Centre Hospitalier Universitaire de Montpellier, France:** V Le Moing, K Klouche

**Centre Hospitalier Universitaire de Lyon, France:** T Ferry, F, Valour

**Centre Hospitalier Universitaire de Nantes, France:** B Gaborit, E Canet, P Le Turnier, D Boutoille

**Centre Hospitalier Universitaire de Reims, France:** F Bani-Sadr

**Centre Hospitalier Universitaire de Rennes, France:** F Benezit, M Revest, C Cameli, A Caro, MJ Ngo Um Tegue, Y Le Tulzo, B Laviolle, F Laine

**Centre Hospitalier Universitaire de Saint-Étienne, France:** G Thiery

**Centre Hospitalier Universitaire de Strasbourg, France:** F Meziani, Y Hansmann, W Oulehri, C Tacquard

**Centre Hospitalier Universitaire de Toulouse, France:** F Vardon-Bounes, B Riu-Poulenc, M Murris-Espin

**Centre Hospitalier Universitaire de Tours, France:** L Bernard, D Garot

**Groupe Hospitalier de Mulhouse Sud Alsace, France:** O Hinschberger

**Hospices Civils de Colmar, France:** M Martinot

**Groupe Hospitalier de Paris Saint Joseph, Paris, France:** C Bruel, B Pilmis

**Hôpital Avicenne, Assistance Publique – Hôpitaux de Paris, France:** O Bouchaud

**Centre Hospitalier Universitaire de Nîmes, France:** P Loubet, C Roger

**Hôpital Bicêtre, Assistance Publique – Hôpitaux de Paris, France:** X Monnet, S Figueiredo

**Hôpital Bichat - Claude Bernard, Assistance Publique – Hôpitaux de Paris, France:** V Godard

**Hôpital Cochin, Assistance Publique – Hôpitaux de Paris, France:** JP Mira, M Lachatre, S Kerneis

**Hôpital Delafontaine, Saint-Denis, France:** J Aboab, N Sayre, F Crockett

**Hôpital Européen Georges-Pompidou, Assistance Publique – Hôpitaux de Paris, France:** D Lebeaux, A Buffet, JL Diehl, A Fayol, JS Hulot, M Livrozet

**Hôpital Henri-Mondor, Assistance Publique – Hôpitaux de Paris, France:** A Mekontso-Dessap

**Hôpital d'Instruction des Armées Bégin, Saint Mandé, France:** C Ficko

**Hôpital Marie Lannelongue, Le Plessis Robinson, France:** F Stefan, J Le Pavec

**Hôpital de la Pitié-Salpêtrière, Assistance Publique – Hôpitaux de Paris, France:** J Mayaux

**Hôpital Saint-Antoine, Assistance Publique – Hôpitaux de Paris, France:** H Ait-Oufella

**Hôpital Saint-Louis, Assistance Publique – Hôpitaux de Paris, France:** JM Molina

**Hôpital Tenon, Assistance Publique – Hôpitaux de Paris, France:** G Pialoux, M Fartoukh

**Hospices Civils de Lyon, France:** J Textoris

**Inserm U1018, Université Paris Saclay, CESP, Paris, France:** M Brossard, A Essat

**Inserm US19-Sc10, Université Paris Saclay, Villejuif, France:** E Netzer, Y Riault, M Ghislain

**Sorbonne Université, INSERM U1136, Institut Pierre Louis d'Épidémiologie et de Santé Publique (IPLESP), Paris, France:** L Beniguel, M Genin, L Gouichiche

**CMG Inserm U1219, Bordeaux Population Health, Université de Bordeaux, Bordeaux, France:** L Moinot, C Betard, L Wittkop

**Cliniques Universitaires de Saint Luc, Bruxelles, Belgique:** L Belkhir

**Centre Hospitalier Régional de la Citadelle, Liège, Belgique:** A Altdorfer, V Fraipont

**Centro Hospital Universitário de Lisboa Norte, Hospital de Santa Maria, Portugal:** S Braz, JM Ferreira Ribeiro

**Centro Hospitalar Universitário São João de Porto, Portugal:** JA Paiva, R Roncon Alburqueque

**Hôpitaux Robert Schuman, Luxembourg:** M Berna

**Luxembourg Institute of Health, Strassen, Luxembourg:** M Alexandre

**Kepler Universitätsklinikum Linz, Linz, Austria:** B Lamprecht

**Paracelsus Medical University Salzburg, SCRI-CCCIT and AGMT, Austria:** A Egle, R Greil

**AGMT Arbeitsgemeinschaft Medikamentöse Tumortherapie, Salzburg, Austria:** R Greil

**Medizinische Universität Innsbruck, Innsbruck, Austria:** M Joannidis

**Acknowledgements**

We gratefully acknowledge the members of the Data Safety Monitoring Board: Stefano VELLA (chair), Guanhua DU, Donata MEDAGLINI, Mike JACOBS, Sylvie VAN DER WERF, Stuart POCOCK, Phaik Yeong CHEAH, Karen BARNES, Patrick YENI, and the independent statistician Tim COLLIER.

We gratefully acknowledge the chief executive officer of Inserm: Dr. Gilles BLOCH.

We acknowledge the RENARCI network: Marion NORET, Pierre TATTEVIN and Albert SOTTO.

We acknowledge the outstanding support of the Clinical Investigation Centers and the Data management and Methodological center: Pierre-Olivier GIRODET (CIC1401, Inserm, CHU Bordeaux), Dominique DEPLANQUE (CIC1403, Inserm, CHRU LIlle), Jean-Luc CRACOWSKI (CIC1406, Inserm, CHU Grenoble), Michel OVIZE (CIC1407, Inserm, Hospices Civil de Lyon), Bernard TARDY (CIC1408, Inserm, CHU Saint-Etinne), Eric RENARD (CIC1411, Inserm, CHU Montpellier), Jean-Noël TROCHU (CIC1413, Inserm, CHU Nantes), Bruno LAVIOLLE (CIC1414, Inserm, CHU Rennes), Wissam EL HAGE (CIC1415, Inserm, CHU Tours), Odile LAUNAY (CIC1417, Inserm, APHP Cochin), Jean-Sébastien HULOT (CIC1418, Inserm, APHP HEGP), Jean-Christophe CORVOL (CIC1422, Inserm, APHP Pitié-Salpétriêre), Mathieu NACHER (CIC1424, Inserm, CH André Rosemon), André CABIE (CIC1424, Inserm, CHU de la Martinique), Xavier Duval (CIC1425, Inserm, APHP Bichat), Philippe LE CORVOISIER Philippe (CIC1430, Inserm, APHP Mondor), Emmanuel HAFFEN (CIC1431, Inserm, CHU Besançon), Marc BARDOU (CIC1432, Inserm, CHU Dijon), Patrick ROSSIGNOL (CIC1433, Inserm, CHU Nancy), Catherine SCHMIDT-MUTTER (CIC1434, Inserm, CHRU Strasbourg), Olivier RASCOL (CIC1436, Inserm, CHU Toulouse), Laurence Meyer (Inserm SC10-US019 ), Linda Wittkop (CMG Bordeaux Inserm U1219), Lambert Assoumou (CMG Inserm U1136 Pitié Salpêtrière).

We acknowledge the monitoring management team: Lydie Beniguel, Christine Betard, Chloé Birklé, Maud Brossard, Charlotte Cameli, Alain Caro, Asma Essat, Michèle Génin, Mathilde Ghislain, Mélanie Grubner, Axel Levier, Cécile Moins, Emmanuelle Netzer, Marie-José Ngo Um Tégué, Isabelle Pacaud and Yoann Riault.

We acknowledge the Clinical research associates: *Belgium*: Nathalie Van Sante, Zineb Khalil; *France*: Malek Ait Djoudi, Lydie Antoine, Christelle Back, Marcellin Bellonet, Assia Benlakhryfa, Nour Boudjoghra, Fabrice Bouhet, Isabelle Calmont, Sabine Camara, Asma Cherifi, Camille Collette, Alexandra De Lemos, Marie Diesel, Elodie Donet, Marine Douillet, Caroline Dubois-Gache, Edith Faillet, Volanantenaina Fanomezantsoa, Stéphanie Flasquin, Shervin Fonooni, Euma Fortes Lopes, Isabelle Gaudin, Blandine Gautier, Quentin Gerome, Marion Ghidi, Pauline Ginoux, Lyna Gouichiche, Marie Granjon, Valérie Guerard, Elina Haerrel, Camille Harpon, Morgane Herbele, Lorrie Lafuente, Dominique Lagarde, Audrey Langlois, Aude Le Breton, Stéphanie Lejeune, Hend Madiot, Bercelin Maniangou, Eric Marquis, Anne-Sophie Martineau, Murielle Mejane, Béatrice Mizejewski, Victoria Mouanga, Brigitte Mugnier, Issraa Osman, Maxence Passageon, Manon Pelkowski, Véronique Pelonde-Erimée, Christine Pintaric, Celina Pruvost, Brigitte Risse, Justine Rousseaux, Christine Schiano, Alexandra Seux, Marielle Simon, Marie-Laure Stupien, Sophie Tallon, Jérémy Tobia, Chaima Traika, Solange Tréhoux, Alice Verdier, Adele Wegang-Nzeufo, Rachida Yatimi; *Luxembourg*: Gloria Montanes; *Portugal*: Catarina Madeira

We acknowledge the pharmacists: Laure LALANDE (Hospices Civils de Lyon), Marine AUSSEDAT (Hospices Civils de Lyon), Jennifer	LE GRAND (APHP Bichat, PARIS), Laura KRAMER (APHP Bichat, PARIS), Sylvie BRICE (CHRU Lille), Sayah MEGUIG (CHRU Lille), Laurent FLET (CHU Nantes), Martine TCHING-SIN	(CHU Nantes), Audrey LEHMANN (CHU Grenoble), Sophie Cerana (CHU Grenoble), Mélanie Minovès (CHU Grenoble), Gwenaël MONNIER (CHU Saint Etienne), Guillaume BECKER (CHRU Strasbourg), Anne HUTT-CLAUSS (CHRU Strasbourg), Sylvia WEHRLEN-PUGLIESE (CHU Nice), Carine GHIONDA (CHU Nice), Marie-Christine RIGAULT (CHU Nice), Christelle BOCZEK (CHU Nice), Justine BELLEGARDE (CHU Nice), Audrey CASTET-NICOLAS (CHU Montpellier), Jean GALLOT-LAVALLEE (CHU Montpellier), Sophie JUAREZ (CHU Montpellier), Valérie CARRE (CHU Montpellier), Fanny CHARBONNIER-BEAUPEL(APHP Pitié-Salpêtrière, Carole METZ (APHP Pitié-Salpêtrière), MURIEL CARVALHO VERLINDE (APHP Mondor), ALAKI THIEMELE (APHP Mondor), delphine LE FEBVRE DE NAILLY (APHP Mondor), Magali FARINES RAFFOUL (CH Annecy Genevois), Franck GUERIN (CH Annecy Genevois), Bénédicte BONNARD (CH Annecy Genevois), Corinne PERNOT (CHU Dijon), Amélie CRANSAC (CHU Dijon), Julie JAMBON (CHU Dijon), Julien MORLET (CHU Dijon), Clotilde Le Tiec (APHP Bicêtre), Aurélie Barrail Tran (APHP Bicêtre), Renée Flore ROY EMA (APHP Bicêtre), Grégory Rondelot (CHR Metz Thionville), Julie Bernez (CHR Metz Thionville), Alexia Hermitte (CHR Metz Thionville), Hélène Perraud (CHR Metz Thionville), Julien Voyat (CHR Metz Thionville), Laurence Ferrier (CHR Metz Thionville), Julie Alips (CHR Metz Thionville), Sarah Di-Filippo (CHR Metz Thionville), Florence Beringuer (CHR Metz Thionville), Michel PREVOT (CHRU Nancy), Sophie MORICE (CHRU Nancy), Brigitte SABATIER (APHP HEGP), Dominique ROUSSEAU (APHP HEGP), Marie KROEMER (CHRU Besançon), Julie VARDANEGA (CHRU Besançon), Magalie VIEILLE (CHRU Besançon), Karen GEORGE (GHR Mulhouse Sud Alsace), Marion TISSOT (GHR Mulhouse Sud Alsace), Laetitia ARNOUX (GHR Mulhouse Sud Alsace), Marjorie BRIMBOEUF (GHR Mulhouse Sud Alsace), Philippe BENOIT (CHU Reims), Frédéric EYVRARD (CHU Toulouse), Caroline SORLI (CHU Toulouse), Corinne GUERIN (APHP Cochin), Jeremie	ZERBIT (APHP Cochin), MERIAM JARDIN SUZCS (HPJS), SOPHIE RENET (HPJS), Alix DE CHEVIGNY (HPJS), Anne Daguenel-Nguyen (APHP Saint-Antoine), Clementine Mayala Kanda (APHP Saint-Antoine), Catherine HAMON-BOUER (CHU Rennes), Pascal LE CORRE (CHU Rennes), Valentine JASPART-LE DU (CHU Rennes), Roza	WILSON (CHU Rennes), Sophie BODDAERT (CHU Amiens), Florian SMAGGHE (CHU Amiens), Mickael DELAUNAY (CHU Amiens), Véronique Dubar (CH Tourcoing), Adeline Danielou (CH Tourcoing), Ghezzoul BELLABES (CHU Bordeaux), Salima ALLAF (APHP Tenon), Jean-Baptiste PAIN (APHP Tenon), Nina DURET-AUPY (CHU Tours), Hélène BOURGOIN (CHU Tours), Thierno DIEYE (Hopital Delafontaine), Anais RAZUREL (Hopital Delafontaine), Jean-Louis LAMAIGNERE (CHU Martinique), Aurélie RISAL (CHU Martinique), Amandine SGARIOTO (Hôpital des Armées Begin), Claire DAVOINE (APHP Hôpital Saint-Louis), Vanessa RATHOUIN (Hôpital Avicenne), Blaise NICAISE (CH Andrée Rosemon), Gregory Gaudillot (CHL, Luxembourg), Emmanuelle Andlauer-Vialette (CHL, Luxembourg), Anne OTTO (HRS, Luxembourg), Francoise FRANTZEN (HRS, Luxembourg), Nathalie BART (HRS, Luxembourg), Martin Wolkersdorfer (Landeskrankenhaus, Salzburg), Astrid Vandorpe (Hôpital Erasme, Bruxelles), Axelle Scarnière (Hôpital Erasme, Bruxelles), Kristel Marquez (Hôpital Erasme, Bruxelles), Nathalie Mathonet (Hôpital Erasme, Bruxelles), Robert Rugwiro (Hôpital Erasme, Bruxelles), Jarode Ducrot (Hôpital Erasme, Bruxelles), Florence Vandenberghe (Hôpital Erasme, Bruxelles), Alexa Jonas (CHR Citadelle, Liège), Valentine Lesenfant (CHR Citadelle, Liège), Blaise Delhauteur (CHR Citadelle, Liège), Annick Peters (CHR Citadelle, Liège), Thi Nguyen (CHR Citadelle, Liège), Jean-François Gouders (CHR Citadelle, Liège), Grégory Opeind (CHR Citadelle, Liège), Adeline GAILLET (Cliniques universitaires Saint Luc, Bruxelles), Ana Carrondo (CHU Lisboa Norte, Hospital Santa Maria, Lisboa), Daniela Ramos (CHU Lisboa Norte, Hospital Santa Maria, Lisboa), Vanessa Côdea (CHU Lisboa Norte, Hospital Santa Maria, Lisboa), Raquel Carneiro CHU Lisboa Norte, Hospital Santa Maria, Lisboa), Ana Lima (CHU Lisboa Norte, Hospital Santa Maria, Lisboa), Tatiana Bento (CHU Lisboa Norte, Hospital Santa Maria, Lisboa).

We thank the pharmacologists: Marie-Claude Gagneu (CHU Lyon), Minh Lê and Gilles Peytavin (CHU Bichat), Benjamin Hennart (CHU Lille), Matthieu Grégoire (CHU Nantes), Françoise Stanke (CHU Grenoble), Xavier Delavenne (CHU Saint-Etienne), Véronique Kemmel (CHU Strasbourg), Noël Zahr (CHU Pitié-Salpêtrière), Patrice Muret (CHU Besançon), Jean-Yves Jouzeau and Nicolas Gambier (CHU Nancy), Rodolphe Garraffo (CHU Nice), Juliette Descoeur (CHU Montpellier), Valérie Furlan (CHU Bicêtre), Peggy Gandia (CHU Toulouse), Jean-Marc Tréluyer (CHU Cochin), Florian Lemaître (CHU Rennes), Anne-Sophie Hurtel-Lemaire, Youssef Bennis and Sandra Bodeau (CHU Amiens), Gilles Paintaud and François Darrouzain (CHU Tours).

We acknowledge the following individuals from the French Data Management Team of Discovery:

Drug supply: Johanna GUILLON, Anne-Marie TABURET.

Inserm-Transfert: Pascale AUGÉ, Mireille CARALP, Nathalie DUGAS, Florence CHUNG, Julia LUMBROSO.

Clinical Research Unit CHU Bichat-Claude Bernard: Camille COUFFIGNAL.

We thank all the staff members involved in data monitoring.

We thank the COVID-19 Partners Platform and Hervé Le Nagard for data hosting.

We thank Theradis PHARMA, Cagnes-sur-Mer, France.

This study was conducted with the support of GILEAD, SANOFI, MERCK and ABBVIE that provided the study drugs.

**Supplementary references**

1.	Haybittle JL. Repeated assessment of results in clinical trials of cancer treatment. The British journal of radiology 1971;44:793-7.

2.	Peto R, Pike MC, Armitage P, et al. Design and analysis of randomized clinical trials requiring prolonged observation of each patient. I. Introduction and design. British journal of cancer 1976;34:585-612.

3.	Mehra MR, Desai SS, Ruschitzka F, Patel AN. RETRACTED: Hydroxychloroquine or chloroquine with or without a macrolide for treatment of COVID-19: a multinational registry analysis. Lancet 2020.
